# Sex-specific dissection of adiposity genetics reveals distinct pathways to endometrial cancer risk

**DOI:** 10.64898/2026.03.29.26349665

**Authors:** Kelsie Bouttle, Dylan M Glubb, Jackson Thorp, Nathan Ingold, Tracy A O’Mara

## Abstract

Excess adiposity accounts for up to 60% of endometrial cancer cases, yet the mechanisms linking adiposity to carcinogenesis and the relevance of sex-specific adiposity genetics to disease risk have been largely unexplored. Using genomic structural equation modelling of six adiposity genome-wide association studies (GWAS), we perform the first sex-stratified adiposity common factor GWAS model, combining data from 2 million people. We identified sex differences in adiposity genetic effects and identified a fourfold larger female-specific causal genetic component relative to males. Female adiposity genetics converged on hormone-responsive and oncogenic pathways directly implicated in endometrial carcinogenesis, a specificity confirmed by stronger female adiposity genetic effects on endometrial cancer but not other hormone-related cancers. Cross-trait analysis identified 26 loci jointly associated with female adiposity and endometrial cancer, including 16 previously unreported loci. GWAS-by-subtraction revealed that only 14.1% of the genetic variance in endometrial cancer is shared with adiposity, with the remainder reflecting adiposity-independent mechanisms captured by established endometrial cancer loci. The adiposity-mediated component converged on insulin-leptin adipocyte signalling and on imprinted and pluripotency-associated developmental pathways, linked by shared nodes such as *PTPN11* and *PPARG*. These findings recast the obesity-endometrial cancer relationship from an epidemiological observation into a mechanistically partitioned genetic programme, and underscores the importance of sex-stratified approaches to resolving how adiposity genetics contributes to disease susceptibility.

## Introduction

Endometrial cancer is the most prevalent gynaecological malignancy in developed nations^1^. In 2022, over 420,000 new cases were diagnosed globally, with age-adjusted incidence increased by 132% over the past three decades and mortality rates have risen by 1.5% annually in the last decade ^1–3^. This unfavorable trajectory contrasts with most other malignancies, where outcomes have generally improved; a concern amplified by the coincident increases in population-level obesity.

Obesity is the strongest known modifiable risk factor for endometrial cancer, with excess body weight accounting for up to 60% of cases and a two- to six-fold increase in disease-specific mortality ^4,5^. Although the epidemiological link is well established, the genetic and biological pathways linking excess adiposity to carcinogenesis remain poorly understood ^6–9^. Body mass index (BMI) is traditionally used as a proxy for excess adiposity in genetic studies due to its measurement simplicity and widespread data availability ^10^. However, BMI conflates fat, lean, and bone mass, and fails to capture variation in fat distribution - an aspect of body composition with known sexual dimorphism and distinct metabolic consequences^11^. This limitation is particularly salient for endometrial cancer, where sex hormones regulate both adipose biology and endometrial proliferation, suggesting that female-specific adiposity genetics may be directly relevant to tumour initiation.

To overcome these limitations, we applied genomic structural equation modelling (GenomicSEM) to jointly model six anthropometric traits, deriving sex-specific latent factors that account for the genetic covariance across anthropometric traits. Leveraging the female-specific adiposity factor from GenomicSEM, we integrated endometrial cancer GWAS into a unified framework that combines pleiotropic locus discovery, colocalisation, and mechanistic decomposition ^12,13^. GenomicSEM was used to perform GWAS-by-subtraction and mediation to partition risk into adiposity-mediated and adiposity-independent pathways, resolving convergent and divergent genetic effects ^14,15^. This provides the first systematic genetic dissection of adiposity’s contribution to endometrial cancer risk.

## Results

A full overview of the study workflow is provided in Figure 1.

**Figure 1.**
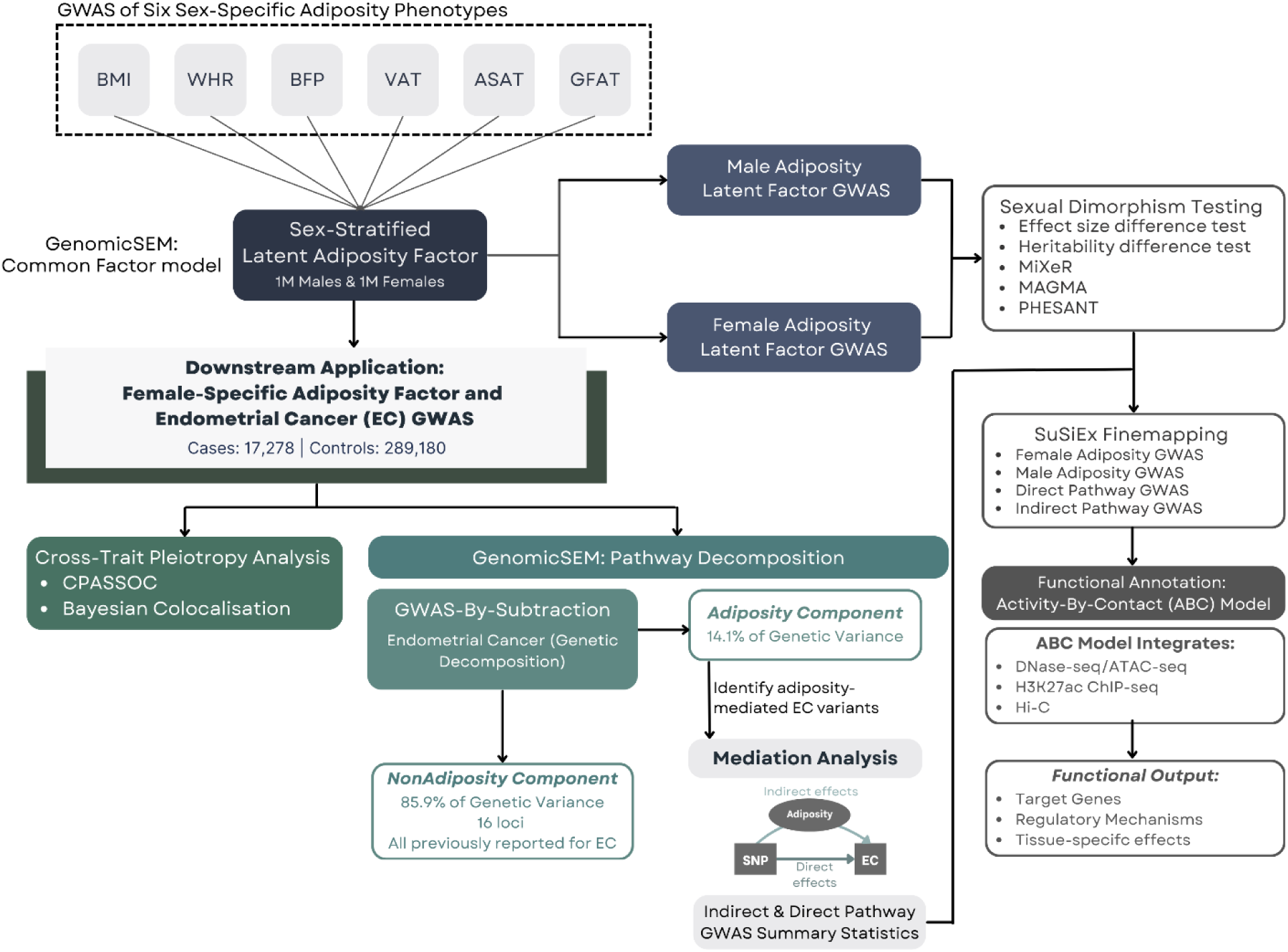
Study overview. BMI, body mass index; WHR, waist-to-hip ratio; BFP, body fat percentage; VAT, visceral adipose tissue; ASAT, abdominal subcutaneous adipose tissue; GFAT, gluteofemoral adipose tissue.

### Sex-stratified genomic structural equation modelling (GenomicSEM) captures distinct adiposity genetics

We used GenomicSEM to derive sex-specific latent adiposity factors from six body composition traits (BMI, WHR, BFP, VAT, ASAT, GFAT) (**Supplementary Table 1**) ^15^. Following quality control of adiposity-associated GWAS, ∼1.17–1.18 million variants were retained for covariance estimation in each sex. LDSC diagnostics showed intercepts near unity and low attenuation ratios across sexes, indicating negligible confounding and supporting robust estimation of the genetic covariance structure. Strong inter-trait correlations were observed among all pairs of adiposity components, supporting a shared genetic basis (**Figure 2a**). After model refinement, both sexes achieved well-fitting latent adiposity factors (female CFI = 0.983, SRMR = 0.058; male CFI = 0.998, SRMR = 0.021), with female-specific factor loadings shown in **Figure 2b** and the corresponding male loadings presented in Supplementary Fig. 1 **(Supplementary Table 2 - 5).**

**Figure 2.**
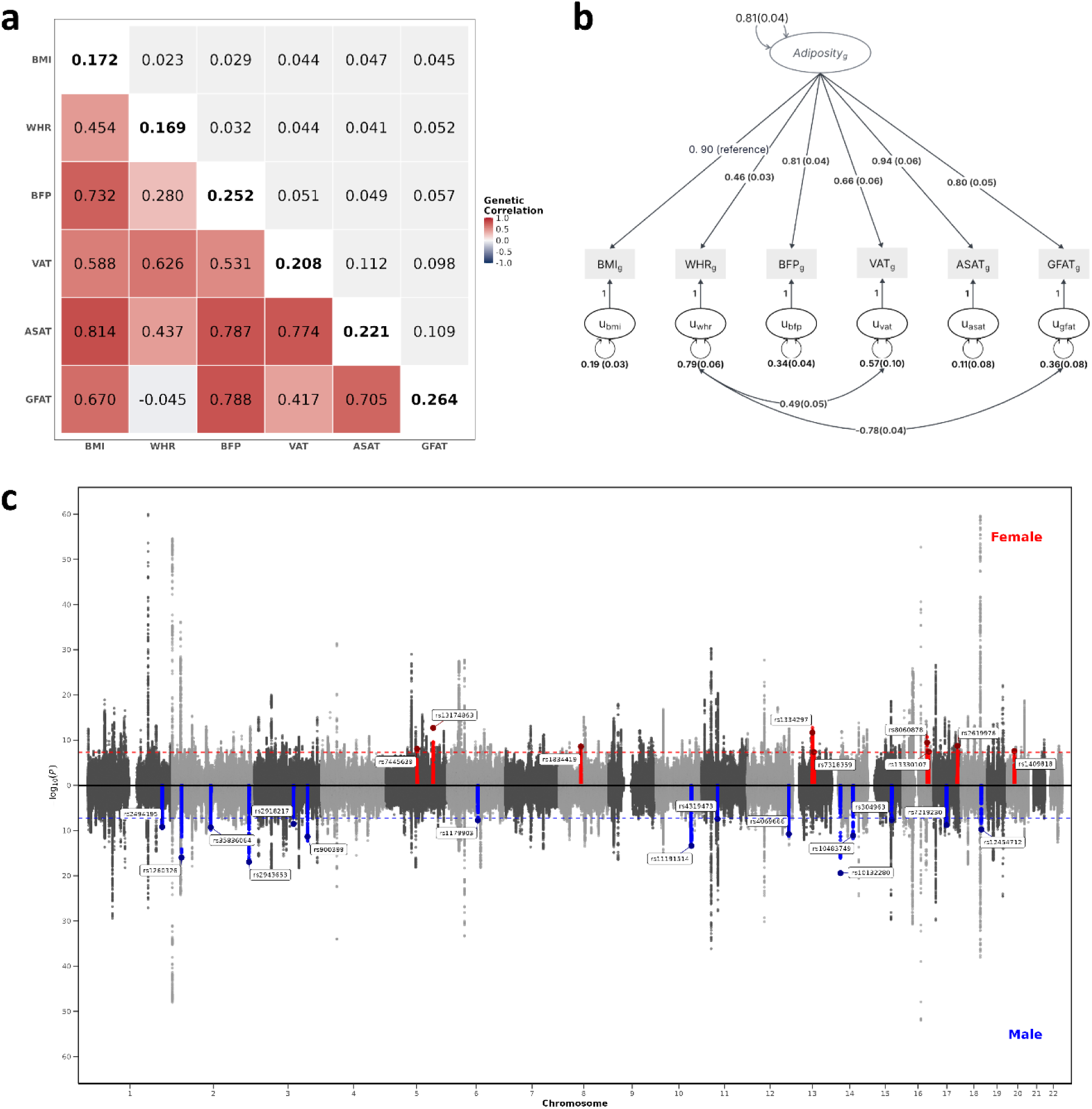
Genetic architecture of sex-specific adiposity: correlations, common factor model, and genome-wide association results. **(a).** Genetic correlations and SNP-based heritability estimates for six adiposity-related traits and the female common factor calculated through linkage disequilibrium score regression. Under and upper off-diagonal elements show genetic correlation coefficients and their standard errors, while diagonal elements display SNP-based heritability values. (**b).** Common factor model displaying standardised loadings derived from GenomicSEM analysis. Residual variances not captured by the latent adiposity factor are indicated by U, where subscript g denotes that the model incorporates genetic covariance relationships among adiposity traits. (**c).** Miami plot showing genome-wide association results for the adiposity common factor in females (top) and males (bottom). Chromosomal position is shown on the x-axis and −log10(P) values are shown on the y-axis. Variants demonstrating significant sex-dimorphic effects are highlighted.

Genome-wide association analyses of the female adiposity factor (N_eff_ = 305,573) identified 262 independent loci, while the male adiposity factor (N_eff_ = 268,683) identified 230 independent loci after LD clumping (**Figure 2c, Supplementary Tables 6 – 8, Supplementary Figures 2 and 3**). This multivariate, sex-stratified approach identified 27 female-specific and 18 loci male-specific loci that had not reached genome-wide significance in the individual contributing sex-specific adiposity GWAS **(Supplementary Tables 9 and 10)**. Q_SNP heterogeneity testing showed that over 90% of loci in both sexes exhibited stable pleiotropic effects across adiposity traits, confirming that the latent factors capture shared adiposity-related genetic architecture rather than trait-specific signals **(Supplementary Note 1; Supplementary Tables 11 and 12; Supplementary Figs. 4 and 5**).

**Figure 3.**
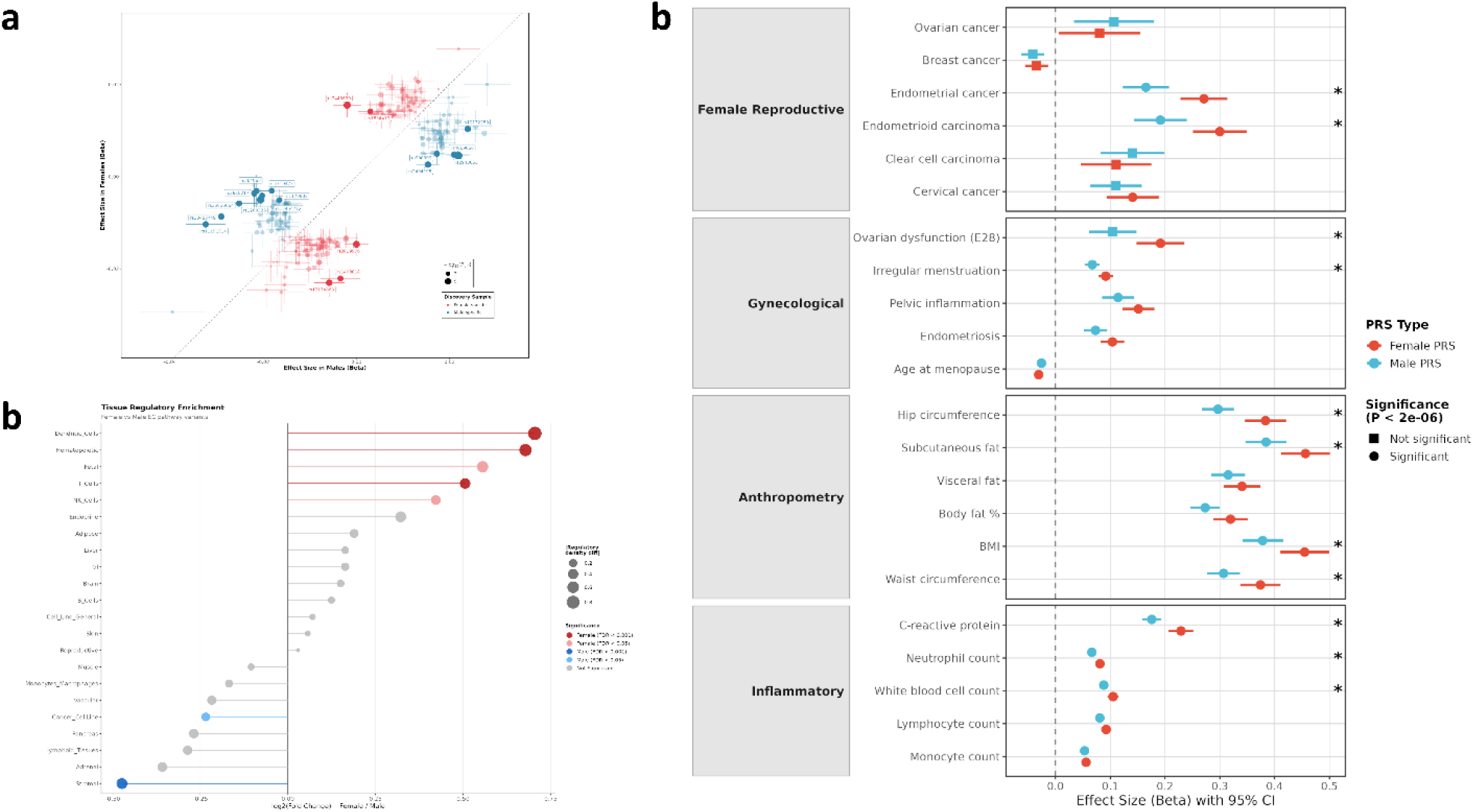
Sex-specific adiposity genetics. **(a).** Male vs female effect size comparison for genome-wide significant lead variants, colored by significance pattern (p < 5×10⁻⁸). The black dashed line indicates perfect correlation (y = x), with male effect sizes on the x-axis and female effect sizes on the y-axis. Red = female-specific significance, blue = male-specific significance. (**b).** Differential tissue regulatory enrichment between female- and male-specific endometrial cancer pathway variants. Lollipop plot showing log2 fold change in gene-to-SNP regulatory density ratio between female- and male-specific variant sets across 22 annotated tissue and cell-type categories. Positive values indicate greater regulatory enrichment in female-specific variants; negative values indicate enrichment in male-specific variants. Point size reflects the absolute difference in regulatory density between variant sets (|Real difference|). Colour denotes statistical significance: dark red, female-enriched FDR < 0.001; pink, female-enriched FDR < 0.05; dark blue, male-enriched FDR < 0.001; light blue, male-enriched FDR < 0.05; grey, not significant. The vertical dashed line indicates log2(Fold Change) = 0. FDR was calculated using the Benjamini–Hochberg method applied to empirical p-values derived from permutation testing. **(c).** Associations with female reproductive health. Forest plot comparing male-derived (blue) and female-derived (red) adiposity polygenic risk scores in combined sex UK Biobank cohort (N=408,780). Circles indicate genome-wide significant associations (P < 2.0 ×10⁻⁶, Bonferroni correction for approximately 25,000 tests); squares indicate non-significant associations. Asterisks (*) denote phenotypes where female and male effect sizes differ significantly (FDR < 0.05) based on Z-tests of beta coefficient differences.

Sex-stratified analyses indicated comparable SNP-heritability and polygenicity, alongside evidence of sex-specific genetic architecture **(Supplementary Note 2)**. The genetic correlation between male and female obesity was high (rg = 0.94, s.e. = 0.03), yet significantly less than one (Z = −2.02, p = 0.044). At the variant level, 11% of adiposity loci displayed significant sex dimorphism after Bonferroni correction (**Figure 3a; Supplementary Table 13; Supplementary Note 2)**. MiXeR modelling confirmed a shared polygenic backbone but identified a non-negligible female-specific set of 390 causal variants, four-fold larger than the male-specific component (90 causal variants; **Supplementary Fig. 6; Supplementary Note 2)**. Gene-based association testing (MAGMA; **Methods**) showed that two-thirds of adiposity genes were sex-specific (386 female-specific, 290 male-specific, 354 shared; **(Supplementary Table 14-17; Supplementary Fig. 7)**. Strikingly, pathway enrichment analysis revealed that female-specific adiposity genes converged on endometrial cancer and Wnt/β-catenin signalling pathways, whereas male-specific genes showed no significant pathway enrichments (FDR < 0.05; **Supplementary Table 18**). To determine whether these divergent genetic architectures operate in distinct tissue contexts, we assigned fine-mapped sex-stratified variants to active regulatory elements across 22 tissue types using ABC enhancer predictions (**Supplementary Note 3**). The sex-stratified variant sets showed similar mapping across tissues, most predominantly to brain, hematopoietic, gastrointestinal, and reproductive tissues, with relatively lower representation for both sets in adipose tissue (**Supplementary Table 19**). However, to determine if the complexity of gene regulation differs within these shared tissue contexts, we compared regulatory density (target genes per variant). Female variants showed higher regulatory density in immune and developmental tissues (e.g. dendritic, hematopoietic and T cells), whereas male variants showed higher regulatory density in stromal tissues (FDR < 0.05; **Figure 3b**; **Supplementary Table 20).**

Phenome-wide association analysis provided an exploratory characterisation of the phenotypic profile associated with sex-specific adiposity genetic liability. Female adiposity genetic effects showed stronger associations than male adiposity genetic effects with endometrial cancer and endometrioid carcinoma (predominantly endometrial cancer), but not with other female hormone-related cancers (P_diff_ FDR < 0.05; **Figure 3c; Supplementary Tables 21-23**). Female adiposity genetic liability was also more strongly associated (P_diff_ FDR < 0.05) with gynaecological (ovarian dysfunction and menstrual irregularity), anthropometric (hip circumference, subcutaneous fat volume and BMI), and inflammatory (neutrophil count, C-reactive protein) traits. In contrast, male adiposity genetic effects showed stronger associations (P_diff_ FDR < 0.05) with insulin use and blood pressure medication, suggesting sex-specific metabolic consequences of adiposity (**Supplementary Table 23).** Together, these findings demonstrate that adiposity genetic architecture diverges between females and males at both the variant and regulatory level, and unstratified adiposity genetics obscures sex-specific disease biology.

### Cross-trait meta-analysis reveals pleiotropic genetic architecture

We applied CPASSOC to test for pleiotropic associations between endometrial cancer and the female-specific adiposity latent factor. This analysis identified 35 genome-wide significant loci (P_CPASSOC ≤ 5 × 10⁻⁸) that also showed at least nominal association with endometrial cancer risk (P_EC < 1 × 10⁻³) (**Figure 4a; Supplementary Tables 24)**. 11 loci showed genome-wide significant associations with endometrial cancer (P_EC < 5 × 10⁻⁸) but no evidence of association with adiposity (P_adiposity > 0.05; **Supplementary Table 25),** indicating adiposity-independent pathways to endometrial cancer risk. Of the remaining 24 loci, five overlap established endometrial cancer GWAS signals (r² > 0.68): rs11263763 at *HNF1B*, rs11066188 at *HECTD4*, rs11065987 at *SH2B3*, rs12602912 at *BPTF* and rs984040 at *HEY2* **(Supplementary Table 26)**. A further three loci showed weaker LD (r² = 0.22–0.35) with known endometrial cancer GWAS risk variants. The remaining 16 loci represent previously unreported endometrial cancer associations. Notably, rs2008514 at *SH2B1*, encoding a key regulator of signalling by the adipokine leptin^16^, demonstrated the strongest evidence of pleiotropy (P_EC = 5.97 × 10⁻⁵; P_CPASSOC = 1.16 × 10⁻²⁸). Complementary CPASSOC analysis with six individual adiposity traits identified 38 loci reaching genome-wide significance (P_CPASSOC ≤ 5 × 10⁻⁸) that also showed at least nominal association with both endometrial cancer (P_EC < 1 × 10⁻³) and the respective adiposity trait (P_trait < 1 × 10⁻³) **(Supplementary Note 4; Supplementary Table 27 – 29).** Of these, 16 overlapped with our adiposity factor analysis, while 10 were subthreshold for either the CPASSOC P-value or did not meet the endometrial cancer significance threshold (P_EC > 1 × 10⁻³), and 12 showed trait-specific associations.

**Figure 4.**
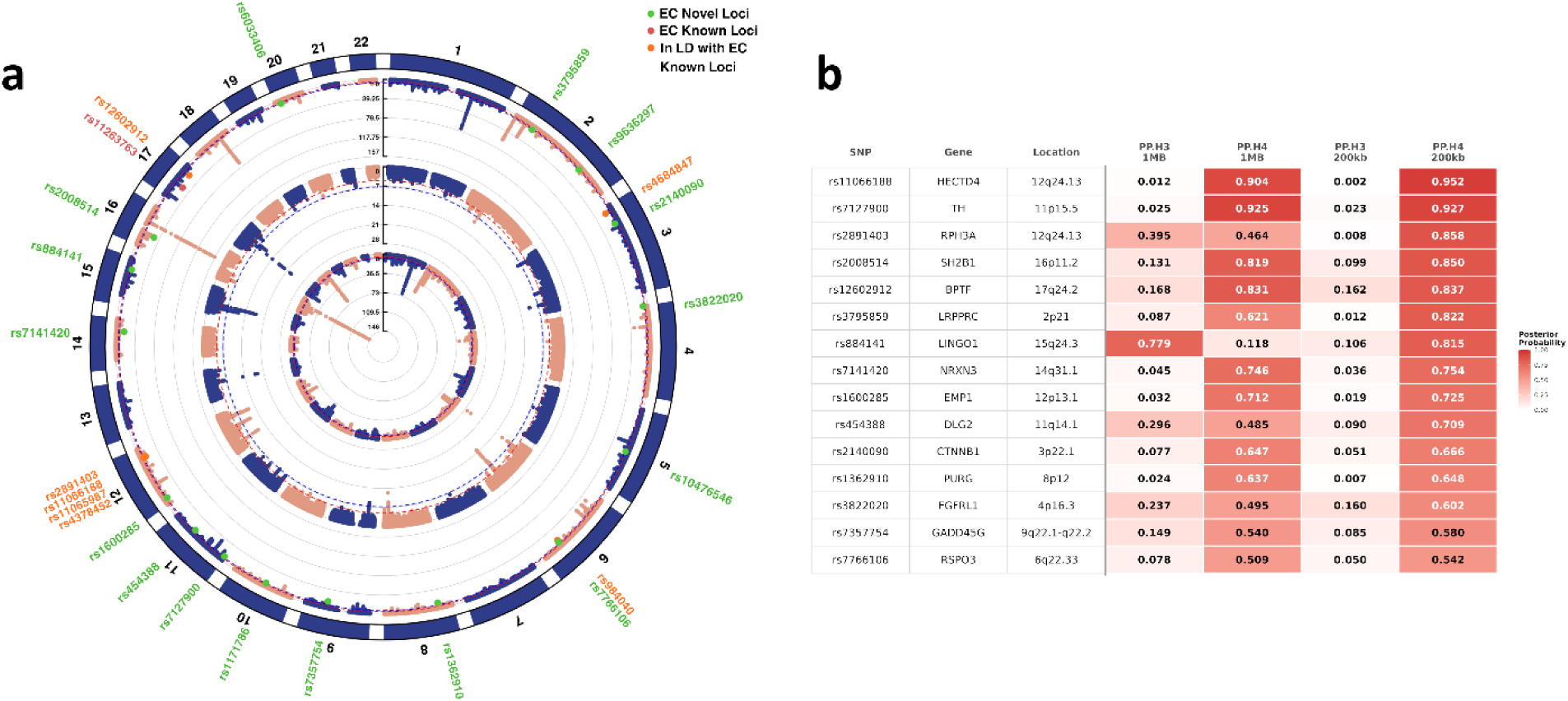
Cross-trait genetic architecture reveals pleiotropic loci between adiposity and endometrial cancer. **(a).** Three concentric circular Manhattan plots display −log10(P) across autosomes (1–22). From outer to inner ring: cross-phenotype association (CPASSOC), endometrial cancer (EC) GWAS, and female-specific adiposity GWAS. Dashed circles denote significance thresholds (blue: P < 5×10⁻⁸; red: P < 1×10⁻⁵). Rim annotations mark variants that are genome-wide significant in CPASSOC and nominally associated in the EC GWAS; green labels indicate putative novel EC loci, red labels indicate previously reported EC loci and orange labels indicate loci in LD with a known EC variant. LD relationships were annotated for interpretation only; no LD threshold was applied for locus definition. Chromosome positions follow the outer ideogram. (**b).** Bayesian colocalisation posterior probabilities for endometrial cancer and female-specific adiposity latent factor. Posterior probabilities for shared (PP.H4) or distinct (PP.H3) causal variants between endometrial cancer and the adiposity latent factor for pleiotropic loci identified by CPASSOC. The lead SNP, nearest gene, and region are annotated on the left. Heatmap cells represent posterior probabilities for PP.H3 and PP.H4 under 1MB and 200KB genomic windows. Loci were included if PP.H4 exceeded 0.5 in at least one window.

**Figure 5.**
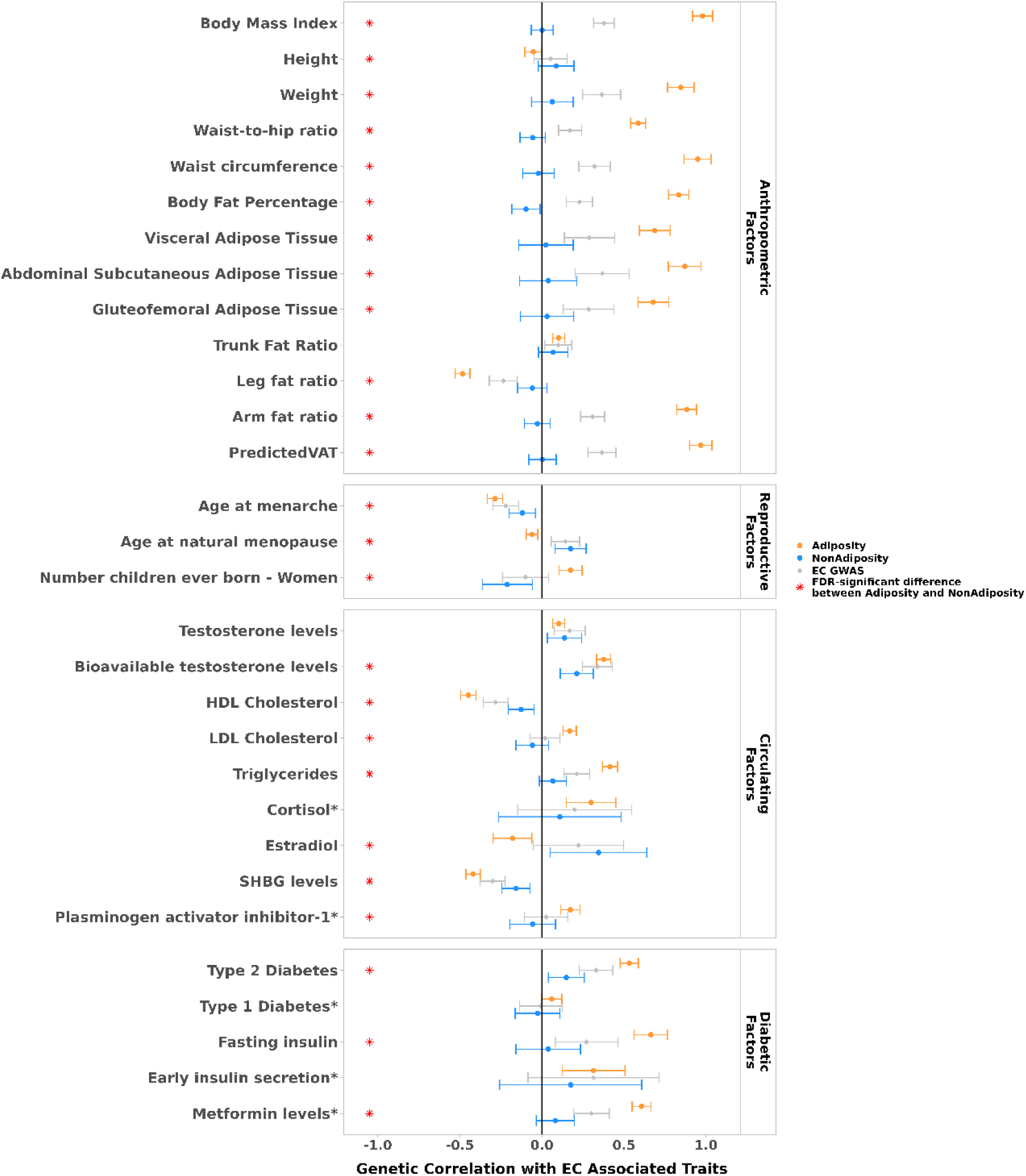
Genetic correlations of *NonAdiposity* and *Adiposity* components with endometrial cancer-associated traits. Dots represent genetic correlations estimated using LDSC. Correlations with Adiposity are shown in orange, NonAdiposity components in blue, and endometrial cancer GWAS correlations in gray. Error bars indicate 95% confidence intervals. Red stars denote statistically significant differences (FDR-corrected P < 0.05, two-tailed test) between adiposity-dependent and adiposity-independent correlations, based on a Z-test that accounts for the standard errors of both estimates. FDR correction was applied across all correlations tested. Traits are grouped by biological categories: anthropometric, reproductive, circulating, diabetic, disease-related, and other traits. Traits marked with an asterisk were only available as combined-sex phenotypes.

### Bayesian colocalisation distinguishes shared from independent causal variants

Among the 24 candidate pleiotropic loci from CPASSOC, four demonstrated strong evidence of shared causal variants between endometrial cancer and the adiposity latent factor (posterior probability for shared variant PP.H4 ≥ 0.75) using 1 Mb windows (**Figure 4b; Supplementary Table 30**). Refining to 200 kb regions increased resolution and identified eight colocalised loci, including four from the broader window. Notably, two independent colocalised signals emerged within 12q24.13.

We also conducted colocalisation for the 38 variants with pleiotropic effects on individual adiposity traits from CPASSOC, which yielded broadly consistent patterns **(Supplementary Note 4)**. Strong evidence was observed at *SH2B1*, *RPH3A*, and *BPTF* (PP.H4 > 0.85), with additional loci showing moderate support (0.50 ≤ PP.H4 ≤ 0.85), suggestive of shared causal variants (**Supplementary Table 29 & 31; Supplementary Figure 8).**

### GWAS-by-subtraction partitions adiposity-associated and adiposity-independent genetic risk

We used GenomicSEM to perform GWAS-by-subtraction, partitioning the genetic architecture of endometrial cancer into components reflecting adiposity-associated (*Adiposity)* and adiposity-independent (*NonAdiposity*) genetic risk (**Supplementary Table 32; Supplementary Figure 9**). These latent variables were modelled as genetically orthogonal (genetic correlation, rg = 0). Sensitivity analyses allowing modest correlations between components (rg = 0.1–0.3) produced consistent results, with endometrial cancer genetic risk predominantly captured by the *NonAdiposity* component **(Supplementary Table 33).**

The *NonAdiposity* component accounted for 85.9% of the total genetic variance in endometrial cancer (**Supplementary Table 32**). Heritability was estimated at *h²* = 0.042 (s.e. = 0.006), derived directly from the model-implied factor variance. The effective sample size (N_eff_) for this component was 3,545. GWAS of this component identified 16 genome-wide significant loci **(Supplementary Table 34; Supplementary Fig. 10)**, all of which have been previously associated with endometrial cancer risk^17,18^. The *Adiposity* component explained 14.1% of the total genetic variance (**Supplementary Table 32**), with heritability estimated at *h²* = 0.143 (s.e. = 0.007). The effective sample size for this component was 310,938. GWAS of this component identified 267 genome-wide significant loci (**Supplementary Table 35; Supplementary Fig. 11)**. None of these variants had reached genome-wide significance in prior endometrial cancer GWAS, although nine showed nominal associations (P < 0.001). Two of these corresponded to loci previously identified as endometrial cancer risk loci: rs12602912 at *BPTF* (r² = 0.71 with lead SNP rs5024718) and rs4684847 at *PPARG* (r² = 0.37 with lead SNP rs11716748)^19^.

We compared genetic correlations of the *Adiposity* and *NonAdiposity* components across 45 anthropometric, reproductive, metabolic, disease, and lifestyle traits, finding significantly different correlations for 31 traits (FDR < 0.05, **Figure 6; Supplementary Tables 36 & 37; Supplementary Figure 12**). The *NonAdiposity* component showed uniformly weak correlations with anthropometric measures (rg = −0.097 to 0.087), whereas the *Adiposity* component demonstrated strong correlations (rg = 0.48 to 0.98), including traits not used to construct the latent factor, such as waist circumference and arm fat ratio. The components diverged across metabolic and hormonal profiles. The *Adiposity* component showed strong correlations with Type 2 Diabetes, fasting insulin, and bioavailable testosterone, and inverse correlations with SHBG and HDL cholesterol, consistent with insulin resistance and hormonal dysregulation. *NonAdiposity* correlations with these metabolic markers were substantially weaker or near zero. Conversely, *NonAdiposity* showed positive correlation with age at menopause (rg = 0.175), whereas the *Adiposity* component showed negligible association (rg = −0.060), suggesting distinct hormonal timing mechanisms.

**Figure 6.**
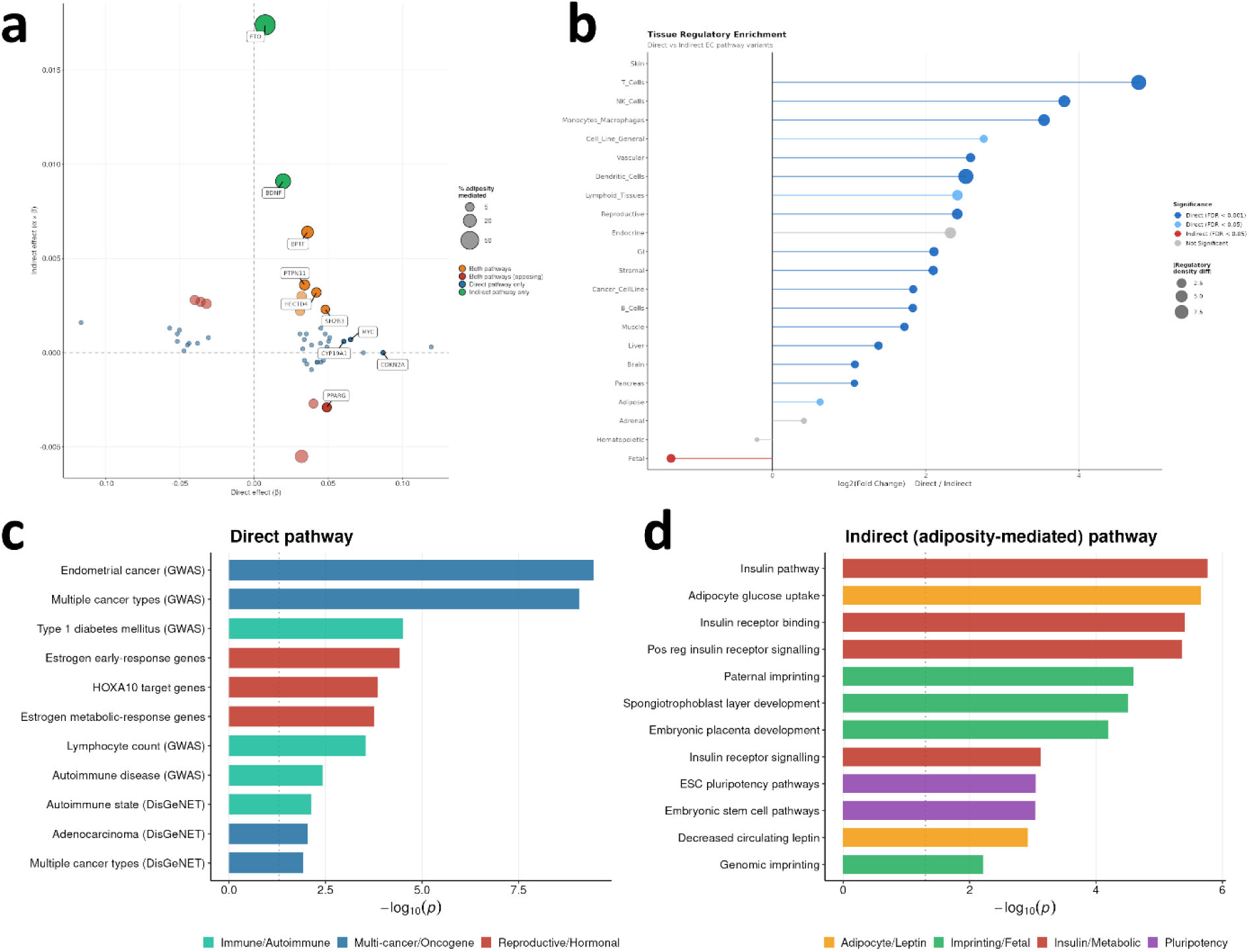
Genetic mediation of endometrial cancer risk through adiposity and tissue-specific regulatory enrichment of pathway variants. **(a).** Scatter plot of SNP-level direct and adiposity-mediated genetic effects on endometrial cancer risk estimated by mediation analysis within the GenomicSEM framework. Each point represents a genetic variant that passed Bonferroni correction (P < 6.92×10⁻⁵). The x-axis shows the direct effect (β, adiposity-independent), and the y-axis shows the indirect effect (α×β, adiposity-mediated). Point size reflects the percentage of the total genetic effect mediated through adiposity. Colour indicates pathway classification: orange, variants with effects through both pathways (concordant); red, variants with effects through both pathways (opposing directions); teal, direct pathway only; green, indirect pathway only. Dashed lines indicate effect = 0. **(b).** Lollipop plot of tissue regulatory enrichment comparing direct (adiposity-independent) versus indirect (adiposity-mediated) pathway variants across 22 annotated tissue and cell-type categories, derived from Activity-By-Contact (ABC) enhancer-gene linking. The x-axis shows log₂ fold change in gene-to-SNP regulatory density ratio (Direct / Indirect); positive values indicate greater enrichment in direct pathway variants. Point size reflects the absolute difference in regulatory density between variant sets. Colour denotes statistical significance: dark blue, direct-enriched FDR < 0.001; light blue, direct-enriched FDR < 0.05; red, indirect-enriched FDR < 0.05; grey, not significant. FDR was calculated by the Benjamini–Hochberg method applied to empirical p-values from permutation testing. **(c, d).** Bar plots showing -log₁₀(p-value) for selected gene sets from ToppFun functional enrichment analysis of ABC target genes linked to (c) direct and (d) indirect (adiposity-mediated) pathway variants. Terms are grouped by functional theme. Direct pathway: Multi-cancer/Oncogene (blue), Reproductive/Hormonal (red), Immune/Autoimmune (teal); Indirect pathway: Insulin/Metabolic (dark red), Imprinting/Fetal (green), Adipocyte/Leptin (orange), Pluripotency (purple). The vertical dotted line indicates p = 0.05. Full results are provided in Supplementary Tables 40–42 and 48–49.

### Mediation analysis apportions pathway-specific genetic effects

We conducted mediation analysis to partition genetic effects on endometrial cancer into direct (adiposity-independent) and indirect (adiposity-mediated) pathways **(Supplementary Figure 13)**. Of 723 nominally associated endometrial cancer variants tested (P < 1×10⁻³; **Supplementary Note 8**, **Supplementary Tables 38–39**), 133 demonstrated significant direct effects independent of adiposity after applying a Bonferroni-corrected threshold (P < 6.91×10⁻⁵, **Supplementary Table 40**). Of these, 16 mapped to previously reported endometrial cancer loci, including *rs7813501* (MYC), *rs1590625* (*CDKN2A*), and rs17601876 (*CYP19A1*). Conversely, 45 variants exhibited significant adiposity-mediated effects **(Supplementary Table 41).** For these variants, adiposity accounted for an average of 12.7% of their total genetic effect on endometrial cancer, indicating that most risk still operates through direct pathways. The strongest indirect effects were observed at rs17817449 (*FTO*; 69.8% mediated), which was the only variant to show a predominantly adiposity-mediated effect, followed by rs962369 (*BDNF*; 31.6% mediated). Four variants at established endometrial cancer risk loci showed modest but significant adiposity-mediated effects: rs6504548 (*BPTF*; 15.2% mediated), rs3184504 (*SH2B3*; 4.6% mediated), rs11066188 (*HECTD4*; 7.1% mediated), and rs13076055 (*PPARG*; 6.3% mediated). Twelve variants demonstrated both direct and adiposity-mediated effects on endometrial cancer risk **(Supplementary Table 42).** Six demonstrated concordant effects where adiposity-mediated and direct pathways reinforced each other (e.g., *SH2B3*, *BPTF* and *PTPN11*), while six exhibited opposing directions where the adiposity-mediated effect modestly counteracted the direct risk (e.g. *PPARG*) (**Figure 6a**).

### Fine-mapping of pathway-specific loci

GWAS analysis of direct (adiposity-independent) and indirect (adiposity-mediated) effect estimates identified 17 and 1,013 independent lead variants, respectively (**Supplementary Tables 43 and 44**). The large number of indirect pathway loci reflects the strong contribution of adiposity genetics; however, only 16 showed nominal association with endometrial cancer (P < 1×10⁻³). Fine-mapping resolved 22 signals across 17 regions for the direct pathway and 32 signals across 16 regions for the adiposity-mediated pathway (**Supplementary Tables 45 and 46**). 9 signals narrowed to single credible causal variants, while others retained broader credible sets with multiple plausible candidates (range 2 – 85 SNPs) **(Supplementary Figure 16**).

To characterise the regulatory context of these distinct effect pathways, we mapped credible causal variants to enhancers across the 22 ABC tissue categories **(Supplementary Note 3)**. Direct pathway variants were significantly more likely to overlap enhancers in cancer cell lines compared to indirect pathway variants (55% vs 32%; FDR = 0.044) and exhibited broad regulatory activity, with higher target gene density across 18 of 22 tissues (FDR < 0.05), including reproductive and all immune cell types (**Figure 6b; Supplementary Table 47**). In contrast, indirect pathway variants showed higher regulatory density only in fetal tissues (3-fold; P = 0.009).

Functional enrichment analysis of ABC target genes reinforced these tissue-specific patterns (FDR < 0.05; **Figure 6c; Supplementary Tables 48-49**). Direct pathway genes were enriched for estrogen response genes and targets of the Müllerian duct regulator *HOXA10*, aligning with high regulatory density in reproductive tissues. Additionally, genes associated with multiple cancer types (e.g. *HNF1B* and *PPARG*), including endometrial cancer, were enriched, mirroring the significant variant overlap observed in cancer cell lines. Direct pathway target genes also showed enrichment for immune cell responses and autoimmune diseases, consistent with the pathway’s heightened immune regulatory density and the pleiotropic immune-metabolic role of *SH2B3*. Adiposity-mediated genes were enriched in imprinted genes and pluripotency signalling pathways, consistent with the increased fetal tissue regulatory density, alongside enrichment in gene sets for multiple insulin signalling pathways, adipocyte traits and circulating leptin levels. These metabolic enrichments were driven by genes including *PPARG* and *PTPN11*, both of which also showed dual adiposity-dependent and independent effects in mediation analysis, reinforcing their roles at the interface of metabolic and tumour-intrinsic risk pathways.

## Discussion

Excess adiposity (obesity) is a well-established risk factor for endometrial cancer, yet the genetic basis of this association and its mechanistic underpinning remain poorly defined. Although males and females exhibit comparable heritability and a largely overlapping polygenic scaffold ^28–32^, we demonstrate that the adiposity-endometrial cancer risk link is, at least partly, attributable to sex-specific genetic architectures. Female adiposity genetics exhibit a fourfold larger causal genetic component than males. Critically, candidate female-specific adiposity genes converge directly on canonical endometrial cancer pathways and Wnt/β-catenin signalling, a pathway mechanistically central to both adiposity^20^ and endometrial carcinogenesis ^21^. These architectural differences extend to regulatory activity, female adiposity variants show higher regulatory density in immune and developmental tissues. PheWAS confirmed that these genetic differences translate into divergent disease risks: female adiposity genetics associated more strongly with endometrial cancer but did not significantly associate with other hormone-related cancers, demonstrating tissue-specific carcinogenic effects. Female genetics also showed stronger effects on gynoid fat distribution (hip circumference, subcutaneous adipose), inflammatory markers, and gynaecological traits. These multi-level distinctions establish that the pathways linking adiposity to endometrial cancer operate through fundamentally sex-specific mechanisms, underscoring the necessity of sex-stratified approaches to resolve causal biology. Together, these observations explain why excess adiposity specifically confers endometrial carcinogenic potential and clarify biological heterogeneity that conventional BMI-based or combined-sex GWAS may obscure^22^.

Cross-trait analyses resolved the shared genetic architecture between female-specific adiposity and endometrial cancer, identifying extensive pleiotropy alongside distinct adiposity-independent loci. Indeed, 11 loci showed genome-wide significant associations with endometrial cancer risk but no evidence of association with adiposity. Among the 24 pleiotropic loci, colocalisation evidence supported shared causal variants at eight; notably, the 12q24.13 region showing two independent colocalised signals, indicating complex regional pleiotropy. Analysis of individual adiposity traits yielded broadly consistent colocalisation patterns. Moreover, a pleiotropic locus at *HNF1B* had variants overlapping with known endometrial cancer risk variants, highlighting the convergence of adiposity and cancer susceptibility on shared hormonal and metabolic drivers. Specifically, *HNF1B* (a key regulator of beta-cell development and insulin secretion)^23^ physically anchors the associations to a carcinogenic event: the promotion of insulin-mediated cellular proliferation ^24^.

GWAS-by-subtraction, using the female-specific adiposity latent factor, partitioned endometrial cancer genetics into *NonAdiposity* and *Adiposity* components. The *NonAdiposity* component captured the vast majority of GWAS-based genetic variance in endometrial cancer (∼86%), recovering 16 genome-wide significant loci, all previously reported endometrial cancer risk loci^17,18^. Genetic correlation analyses reinforced this with the *NonAdiposity* component showing weak correlations with anthropometric traits but positive correlations with reproductive timing and circulating estradiol, patterns consistent with estrogen-driven carcinogenesis. In contrast, the *Adiposity* component explained only ∼14% of genetic variance yet yielded 267 genome-wide significant loci. None of the lead variants had previously reached genome-wide significance in endometrial cancer GWAS, although nine showed nominal associations (P < 0.001), including signals in LD with previously reported risk loci at *BPTF* and *PPARG*. This small, adiposity-associated fraction reconciles the strong epidemiological link between obesity and endometrial cancer with emerging evidence indicating that the obesity-cancer link is largely environmentally forged rather than genetically predetermined ^25^. The *Adiposity* component displayed strong correlations with type 2 diabetes, fasting insulin and reduced SHBG, metabolic hallmarks of insulin resistance and adipokine dysregulation ^26–28^, indicating that this component superimposes an auxiliary metabolic layer of risk.

Consistent with the GWAS-by-subtraction findings, mediation analysis showed that only a small subset of variants act meaningfully through adiposity, and even for these, indirect effects are typically modest. For variants with adiposity-mediated effects, adiposity on average explained only 13% of each variant’s effect on endometrial cancer risk. The locus represented by *FTO*, a canonical obesity gene, was the only locus with a predominant adiposity-mediated effect ^29^. Twelve variants carried both direct and mediated effects, with some, including established endometrial cancer GWAS risk loci at *SH2B3* and *BPTF*, demonstrated reinforcing effects on risk; whereas others, such as that represented by *PPARG*, displayed opposing directions where adiposity-mediated protection partially offset direct risk, highlighting that adiposity can both amplify and buffer tumour-intrinsic susceptibility at specific loci

Fine-mapping and enhancer-variant target gene annotation revealed sharply divergent regulatory architectures for the direct and adiposity-mediated risk pathways. Direct variants were enriched in cancer cell line enhancers, with target genes converging on estrogen-response pathways, Müllerian duct development via HOXA10, and multi-cancer risk genes, consistent with broad tumour-intrinsic regulatory activity. Adiposity-mediated variants showed regulatory enrichment selectively in fetal tissues, with target genes relevantly implicated in insulin and leptin signalling, and in pluripotency-associated developmental programmes. At the nexus of these programmes sits PTPN11, encoding a key signalling molecule involved in mitogenic activation and metabolic control^30^, alongside other shared nodes such as PPARG, a master regulator of adipocyte differentiation and insulin sensitivity ^31^. SH2B1, an adaptor that potentiates leptin signalling^32^ and shows pleiotropic effects on adiposity and endometrial cancer risk in our cross-trait analyses, further links this metabolic circuitry to tumour-intrinsic susceptibility. Leptin, secreted by adipocytes and elevated in obesity, has been independently associated with endometrial cancer risk^33^, and Mendelian randomisation supports a causal role for insulinaemia in endometrioid adenocarcinoma^7^, lending epidemiological support to this genetically defined metabolic axis. Indeed, adiposity-mediated variants, although modest in effect, converge on coherent adipocyte and developmental programmes that highlight biological routes through which environmentally acquired obesity amplifies endometrial cancer risk.

Several limitations merit consideration. First, as our analyses were based on European-ancestry GWAS, the effects linked to the adiposity factor may not generalise to non-European populations. Second, marked sample size disparity between the combined six adiposity-related GWAS and endometrial cancer data may bias discovery toward adiposity loci, particularly in GWAS-by-subtraction and multi-trait CPASSOC analyses. This would complicate differentiation between adiposity and cancer-specific genetic effects. Third, while we carefully balance model complexity and parsimony, residual genetic effects may remain uncaptured due to limitations in measurement quality and data completeness. Fourth, the adiposity-mediated and adiposity-independent pathways are not fully orthogonal, and our GenomicSEM framework provides a statistically simplified representation that may not fully capture complex horizontal pleiotropy. Fifth, our gene-level and pathway inferences rely on MAGMA-based gene association, ABC enhancer–gene linking, and curated pathway databases, which provide probabilistic rather than definitive mappings and are subject to known biases in functional annotation.

This study delivers locus-level resolution and pathway-level coherence, yielding mechanistic insights into the genetic relationship between endometrial cancer risk and adiposity that single-method approaches could have overlooked. Future work will leverage multivariate clustering to disentangle mechanisms in which genetic components of adiposity confer endometrial cancer risk. By stratifying loci and polygenic profiles across estrogen signalling, inflammatory pathways, adipokines, and insulin-related traits, this approach will enable finer resolution of the molecular programs through which adiposity influences tumour susceptibility. Such analyses will shift the field away from treating obesity as a homogeneous exposure and toward identifying specific biologically active forms of adiposity that promote endometrial carcinogenesis. Functional studies in relevant cellular and animal models, prioritising fine-mapped loci and their putative target genes, will be essential to validate causal mechanisms and identify tractable nodes for intervention, including the potential repurposing of existing metabolic therapies for prevention or therapy.

## Conclusion

This work demonstrates that the genetic architecture of excess adiposity is biologically distinct, with female specific adiposity genetics directly interfacing with tumour-intrinsic pathways to shape endometrial cancer susceptibility. By partitioning heritable risk into adiposity-independent pathways and an adiposity-mediated axis, we reveal that a smaller fraction of genetic risk is channelled through adiposity, converging on insulin and leptin signalling and on imprinted and pluripotency-associated developmental pathways, linked by shared nodes such as PTPN11 and PPARG. These findings recast the obesity–endometrial cancer relationship from a statistical association into a mechanistically partitioned genetic programme and underscore the broader importance of sex-stratified approaches to understanding how adiposity genetics shapes disease susceptibility. They establish a platform for pathway-specific risk stratification and for metabolic interventions that directly address carcinogenic adiposity by targeting the same pathways through which environmentally acquired obesity amplifies risk.

## Material and Methods

### Data Summary

This study leveraged the largest genome-wide association study (GWAS) data for endometrial cancer to date, comprising 17,278 cases and 289,180 controls ^19^. Summary-level GWAS data for six adiposity-related traits were sourced from publicly available repositories, comprising up to 1,030,397 female and 906,579 male participants of European ancestry (**Supplementary Table 1)**. All genomic coordinates were standardised to GRCh37/hg19 genome-build where necessary. For cross-phenotype analysis, mtCOJO was used to adjust data for BMI where BMI-adjusted summary statistics were not already available, using linkage disequilibrium (LD) estimated from the 1000 Genomes Phase 3 European reference panel ^34^.

### Latent Adiposity Factor GWAS

We performed genome-wide association analyses using Genomic Structural Equation Modelling (GenomicSEM) v0.0.5, a two-stage approach that models genetic relationships between traits using GWAS summary statistics ^15^. This method accounts for sample overlap through LD score regression-based genetic covariance estimation and facilitates the integration of data from multiple sources. First, multivariable LD score (LDSC) regression was used to estimate the genetic covariance matrix (and its associated sampling covariance matrix) for the set of traits. Second, a structural equation model was specified and model parameters estimated by minimising the difference between the model-implied and empirical covariance matrices. Models were estimated using a diagonally weighted least squares (DWLS) estimator.

Model fit was assessed using the standardised root mean square residual (SRMR), Akaike Information Criterion (AIC), and Comparative Fit Index (CFI), with residual covariances added only until satisfactory fit was achieved ^15^. Where initial model fit was unsatisfactory, we examined the residual matrix, defined as the difference between the observed genetic correlation matrix and that implied by the factor model, to identify trait pairs whose genetic covariance was not well captured by the common factor alone. Residual covariances were added to the model sequentially, prioritised by magnitude, and the model was refit at each step. This process was repeated until good model fit was achieved, with the number of residual covariances kept to the minimum necessary to avoid model saturation.

To investigate the shared genetic architecture underlying adiposity as a latent trait, we applied GenomicSEM to estimate a common factor model for female-specific and male-specific adiposity using the six adiposity-related traits: BMI, waist-hip ratio (WHR), body fat percentage (BFP), visceral adipose tissue (VAT), abdominal subcutaneous adipose tissue (ASAT), and gluteofemoral adipose tissue (GFAT) (**Supplementary Table 1**). The model was specified using unit variance identification. We then performed a GWAS of the latent adiposity factor using the *commonfactorGWAS* function in Genomic SEM. Genetic variants were retained if present in all six GWAS datasets, the 1000 Genomes Phase 3 reference panel, and had a minor allele frequency (MAF) > 0.005. This yielded 6,652,069 genetic variants for males and 7,073,715 for females.

Latent adiposity factor GWAS results for each sex were LD clumped using r^2^ > 0.001 and a 500 kb window to identify independent lead genetic variants at each loci reaching genome-wide significance (P < 5 × 10^-8^), using PLINK v1.9 ^35^ and a 1000 Genomes LD reference panel of European descent (phase 3v5)^36^. The effective sample size of each adiposity common factor GWAS was calculated as per Mallard et al (2022) ^37^.

### Fine-mapping of lead Genetic Variants

Statistical fine-mapping was performed using SuSiEx to identify putative causal variants underlying genome-wide significant associations from male- and female-specific adiposity summary statistics ^38^. Independent lead variants were first identified by clumping summary statistics in PLINK v1.9 ^35^. To account for LD, chromosome-specific LD matrices were constructed from the UK Biobank 2018 European ancestry reference panel. For each genome-wide significant lead SNP (P < 5×10⁻⁸), ±500 kb regions were analysed to define 95% credible sets, permitting up to 10 causal signals per region.

### Sexual Dimorphism Lead Variant Effect Size Comparison

We selected genetic variants for sex dimorphism testing based on the following criteria: genome-wide significant association (P < 5×10⁻⁸) in one sex and no genome-wide significant association (P > 5×10⁻⁸) in the opposite sex within ±500 kb of the lead SNP. This yielded 130 female-specific and 98 male-specific genetic variants. We tested for sex-dimorphic effects in R (v4.5.0): using the t-statistic:

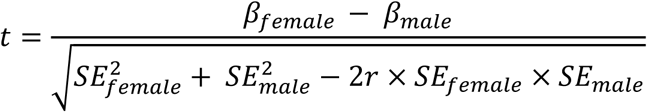

Where β represents the effect estimate of the genetic variant in each sex, SE is the standard error of the effect estimate, and r is the genome-wide Spearman rank correlation coefficient between SNP effect estimates in females and males (r = 0.323). We calculated two-tailed P-values (P_diff_) from the t-distribution. We applied Bonferroni correction for multiple testing across 228 tests (0.05/228 = 2.19×10⁻⁴). Genetic variants were classified as having stronger effects in females or males based on the absolute magnitude of sex-specific effect estimates, considering direction of effect and significance levels when effects were in opposite directions, as per Pulit et al. (2019)^39^.

### Sex Differences in SNP-Based Heritability

To test whether overall SNP-based heritability (h²) estimates differed between sexes, we calculated a z-score:

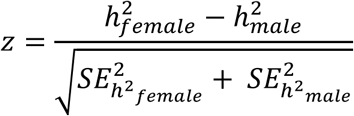

where h² represents the SNP-based heritability estimate and SE_h²_ is its standard error. Z-scores were converted to two-tailed P-values using the standard normal distribution:

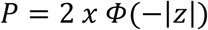

### MiXeR

We used MiXeR v1.3 to characterise the genetic architecture of adiposity in sex-stratified GWAS datasets and to quantify polygenic overlap between sexes ^40^. MiXeR models SNP effects as a Gaussian mixture of null and causal components, allowing estimation of the subset of variants contributing to trait variance. Univariate MiXeR estimated polygenicity (π), SNP-heritability (h²SNP), and the number of causal variants explaining 90% of h² SNP for each sex, which prevents extrapolation into negligible-effect regions. Bivariate MiXeR quantified the proportion of female-specific, male-specific, and shared causal variants and the genetic correlation (rg) between sexes. All models incorporated LD derived from the 1000 Genomes Phase 3 European reference panel^36^.

### MAGMA

Gene-based association tests were conducted using Multi-marker Analysis of GenoMic Annotation (MAGMA) v1.08 to aggregate SNP-level GWAS signals into gene-level statistics for female and male adiposity GWAS separately^41,42^. SNPs were mapped to 18,403 protein-coding genes (NCBI build 37.3), extending 35 kb upstream and 10 kb downstream. MAGMA’s SNP-wise model was applied using LD patterns from the 1000 Genomes Project Phase 3 European reference panel ^36^. Bonferroni correction set the significance threshold at P < 2.72×10⁻⁶.

### Pathway Analysis

Significant genes from MAGMA were included in a pathway enrichment analysis using ToppFun (ToppGene Suite; https://toppgene.cchmc.org/)^41,42^. Enrichment was performed separately for each set of sex-specific genes across KEGG, Reactome, WikiPathways and Reactome pathways databases. Statistical enrichment was evaluated using hypergeometric testing with Benjamini–Hochberg false discovery rate (FDR) correction, and terms with FDR-adjusted P < 0.05 were considered significant.

### Activity-By-Contact (ABC) Model

We used the Activity-By-Contact (ABC) Enhancer-Gene dataset v3 model to map sex-stratified adiposity SNPs to putative target genes across 131 cell or tissue samples ^43^. ABC integrates chromatin accessibility (DNase-seq, H3K27ac histone marking, and 3D contact frequency (Hi-C)) to predict enhancer-promoter interactions. Variants overlapping ABC-predicted enhancers (ABC score > 0.015) were assigned to their predicted target genes. ABC biosamples were aggregated into 22 functional tissue categories **(Supplementary Note 3)**.

Tissue enrichment for each sex-stratified variant set was assessed by calculating the percentage of credible SNPs overlapping ABC enhancers in each tissue category. Individual SNPs were mapped to multiple tissues. Regulatory density (target genes per variant) was calculated for each tissue as the ratio of unique target genes to unique credible SNPs mapping to that tissue. Sex-specific differences in regulatory density were tested via permutation (n=10,000): for each permutation, sex labels were randomly shuffled across the credible SNP set, and the difference in regulatory density (Female ratio – Male ratio) was recalculated. Empirical two-sided p-values were computed as the proportion of permutations where |permuted difference| ≥ |observed difference|. FDR correction was applied across all 22 tissues using the Benjamini-Hochberg method (FDR < 0.05). Analysis code is available at https://github.com/glubbster/obesity-abc-permutation.

### PHEnome Scan ANalysis Tool (PHESANT)

Sex-stratified adiposity polygenic risk scores (PRS) were derived using LDpred2-auto with Genomic SEM-derived summary statistics from males and females ^44,45^. LDpred2-auto estimated posterior SNP effect sizes while accounting for LD structure, producing optimised SNP weights for each sex independently. Individual-level PRS were calculated by applying these sex-specific weights to UK Biobank genotype data using PLINK v2.0, with scores computed separately per chromosome (1-22) and summed to generate genome-wide PRS. Within each sex, PRS were standardised prior to downstream analyses.

Phenome-wide association analyses (PheWAS) were conducted using PHESANT to test associations between sex-stratified adiposity PRS and approximately 25,000 phenotypes ^44,45^. Analyses were performed using both female-derived and male-derived PRS, each evaluated across the full UK Biobank sample. Because the adiposity GWAS summary statistics used to derive PRS weights likely to include UK Biobank participants, these analyses were not fully independent; therefore, results should be interpreted as descriptive and hypothesis-generating, providing phenotypic characterisation and construct validation rather than independent replication.

Associations were evaluated using linear regression for continuous outcomes and logistic regression for binary outcomes, with all models adjusted for age, age², assessment centre, genotyping array, and the first 10 genetic principal components. A Bonferroni-corrected significance threshold of P < 2.0 × 10⁻⁶ (0.05/25,017 tests) was applied.

### Cross-trait meta-analysis

To identify shared genetic loci between endometrial cancer and adiposity, we performed cross-trait genome-wide association analyses at the variant level using CPASSOC, applying the framework to both the female adiposity latent factor and the six individual BMI-adjusted adiposity traits **(Supplementary Table 1)** ^12^. CPASSOC combines GWAS summary statistics across correlated phenotypes to detect pleiotropic loci while accounting for population stratification and cryptic relatedness. It provides two test statistics: S_Hom_ (assuming homogeneous shared effect directions across traits), and S_Het_ (allowing for heterogeneity in shared effect sizes and directions). We used S_Het_ for all analyses.

We performed LD clumping of the CPASSOC results to identify lead genetic variants using PLINK v1.9. We defined a variant a pleiotropic if it reached genome-wide significance in the cross-trait analysis (P_CPASSOC < 5 × 10⁻⁸) and showed supporting evidence of association with endometrial cancer (P < 0.001).

### Colocalisation

Colocalisation analysis implemented in the *coloc* R package (v5.2.3) ^13^ was used to assess whether the pleiotropic genetic variants identified in the CPASSOC analysis share a causal variant between traits. For each locus, *Coloc* computes approximate Bayes factors and estimates posterior probabilities for five hypotheses:

- H0: No association with either trait
- H1: Association with trait 1 only
- H2: Association with trait 2 only
- H3: Association with both traits, due to two independent variants
- H4: Association with both traits, as a result of one shared variant

For each lead variant identified in CPASSOC, we used summary statistics for the respective adiposity-related traits to calculate posterior probabilities of a shared variant with endometrial cancer. This was done using default prior settings and in a ±500 kb region of the lead SNP and using a narrower ±100 kb window to assess the robustness of the results to region size. A region was considered to show evidence of colocalisation if the probability supporting a shared causal variant (PP.H4) was greater than 0.75.

### GWAS-by-subtraction

We partitioned adiposity-independent genetic effects on endometrial cancer using GWAS-by-subtraction implemented in GenomicSEM ^14^. Summary statistics from the endometrial cancer GWAS and the female-specific adiposity factor were incorporated into a multivariate GenomicSEM model **(Supplementary Fig 9; Supplementary Table 1).** The endometrial cancer phenotype and the adiposity factor were initially regressed onto a latent variable representing adiposity regulation (*Adiposity*). Subsequently, endometrial cancer was regressed onto a second latent factor capturing residual genetic variance independent of adiposity (*NonAdiposity*). Residual variance of endometrial cancer was fixed to zero, ensuring endometrial cancer is fully explained by these two components. Genetic decomposition separated endometrial SNP effects into two orthogonal pathways: those shared with adiposity and those independent of it. GWAS analyses were then performed separately for *Adiposity* and *NonAdiposity* components, yielding summary statistics reflective of the two distinct pathways to endometrial cancer susceptibility.

GWAS summary statistics were LD clumped using PLINK v1.9. Effective sample size was derived as recommended by Demange et al. (2021)^14,37^. LDSC was used to compare *Adiposity* and *NonAdiposity* components previously associated with endometrial cancer-associated phenotypes. These traits were grouped into six themes: anthropometric, reproductive, circulating factors, diabetic factors, disease outcomes, and lifestyle risk factors.

### Mediation

To further characterise variant-specific mechanisms linking adiposity to endometrial cancer, we conducted mediation analysis within the GenomicSEM framework. Whereas GWAS-by-subtraction partitions genome-wide genetic variance into adiposity-associated and adiposity-independent components, mediation analysis evaluates individual SNP effects by decomposing them into direct and adiposity-mediated pathways. Mediation analysis was implemented using the *userGWAS()* function. For each SNP, we estimated: the direct effect (c) representing the SNP effect on endometrial cancer independent of adiposity; the indirect effect (a×b) representing the product of SNP effect on adiposity (a) and adiposity effect on endometrial cancer (b). The total effect is expressed as: Total Effect = Direct Effect (c) + Indirect Effect (a×b), and Proportion Mediated = Indirect Effect (a×b) / Total Effect (c+(a×b)) (**Supplementary Fig 13**). The adiposity factor derived from our previous common factor analysis served as the mediating variable in all models.

A total of 758 independent variants reaching nominal significance in the endometrial cancer GWAS (P < 0.001) were prioritised for mediation testing to capture variants with evidence of association with endometrial cancer and evaluate whether their effects operate through adiposity-mediated pathways. Of these, 35 variants were unavailable in both datasets and no suitable proxies could be identified, resulting in 723 variants included in the analysis (Supplementary Note 5). To account for multiple testing, we applied Bonferroni correction yielding a significance threshold of P < 6.92×10⁻⁵ (0.05/723).

Genome-wide significant loci identified in the pathway-specific GWAS summary statistics generated from the mediation analysis were prioritised for fine-mapping to resolve putative causal variants underlying adiposity-mediated (indirect) and adiposity-independent (direct) effects. Variants from each pathway were clumped using PLINK v1.9 ^35^. For the indirect pathway, variants were retained if they showed supporting evidence of association with endometrial cancer (P < 0.001). Fine-mapping was then performed using SuSiEx as described above.

## Data Availability

Sex-stratified adiposity GWAS summary statistics generated in this study will be available at: https://zenodo.org/records/19124077?token=eyJhbGciOiJIUzUxMiJ9.eyJpZCI6IjAyNDk5NTYzLTY5MjktNDJjYi04ODZlLTM5NTU1ZWZhZGUzYSIsImRhdGEiOnt9LCJyYW5kb20iOiI1ODI1NDIyMmQwMWEwZTk5NTk3ODYzMzkwOTdmMDk0OCJ9.jwf9dQV_tRiINwA-XFKRLsoenBNQBvFCC_DauAsBNMBKXLZAIpNKnzA6pBbrHvLDlxfHhuUCy6tvrHRbtmLGtg Under embargo until publication

## Acknowledgements

We thank the research participants for making this study possible. This work was conducted using the UK Biobank Resource (application number 25331). We thank the GWAS Catalog for storing and facilitating access to publicly available GWAS summary statistics. We thank the QIMR Berghofer Genome Informatics team for maintaining the high-performance computing cluster and providing computational advice.

This work was supported by a National Health and Medical Research Council (NHMRC) of Australia Investigator Grants (APP1173170 and #ID 2041819) awarded to T.A.O’M, and an NHMRC Ideas Grant (#ID 2036849) awarded to T.A.O’M, D.M.G. and N.I. J.T. is supported by an NHMRC Investigator Grant EL1 (#ID 2027002). K.B. is supported by a QIMR Berghofer Cancer Genetic Susceptibility PhD Scholarship and a University of Queensland Research Training Scholarship. The funders had no role in study design, data collection and analysis, interpretation of results, or manuscript preparation.

## Notes

### Competing Interest Statement

The authors have declared no competing interest.

### Author Declarations

The UK Biobank. The GWAS Catalog

