## Supplementary Notes + Figures for "Sex-specific dissection of adiposity genetics reveals distinct pathways to endometrial cancer risk"

Supplementary Figure 10. Manhattan Plot of Non-Adiposity Component from GWAS-by-Subtraction Analysis .......... 1

Supplementary Figure 11.  Manhattan Plot of Adiposity Component from GWAS-by-Subtraction Analysis .......... 2

**Supplementary Figures**

**Supplementary Figure 1. Male Multivariate Adiposity Common Factor GenomicSEM Model**

Common factor model displaying standardised loadings derived from Genomic SEM analysis. Residual variances not captured by the latent adiposity factor are indicated by U, where subscript g denotes that the model incorporates genetic covariance relationships among adiposity traits.

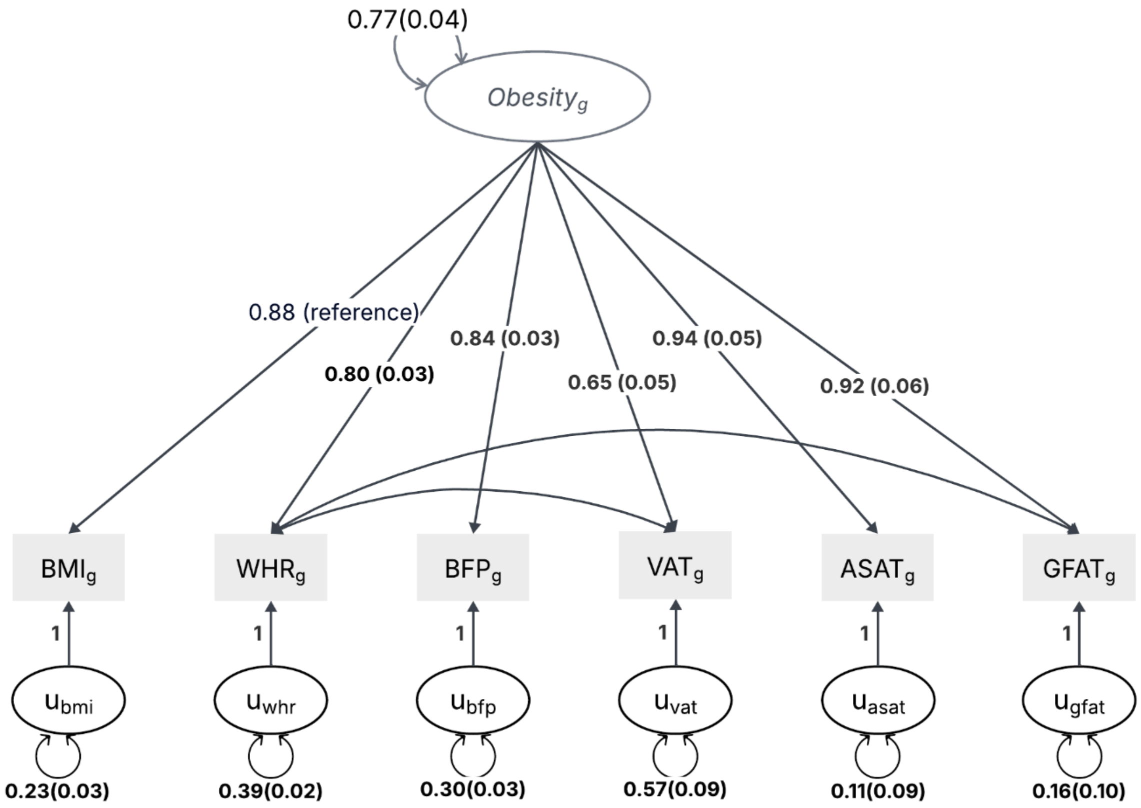

**Supplementary Figure 2. Manhattan and Quantile-Quantile (QQ) plot for Multivariate Female Adiposity GWAS.**

**a**, Manhattan plot showing GWAS results for the female adiposity factor plotted by chromosomal location. Chromosomal position is shown on the x-axis, with the y-axis displaying −log10(P) values from association tests between SNPs and the latent adiposity factor. **b,** Quantile–quantile (QQ) plot comparing observed versus expected −log10(P) under the null for the same GWAS.

**a**

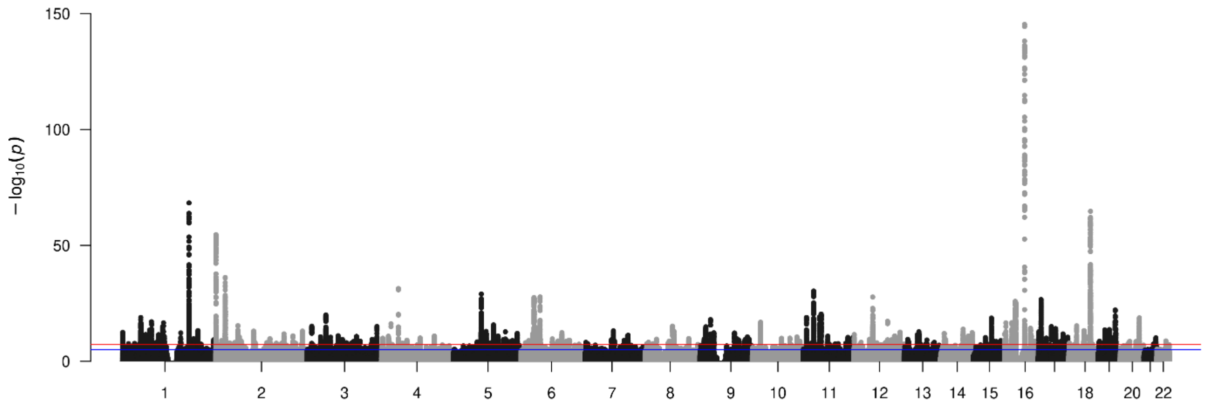

**b**

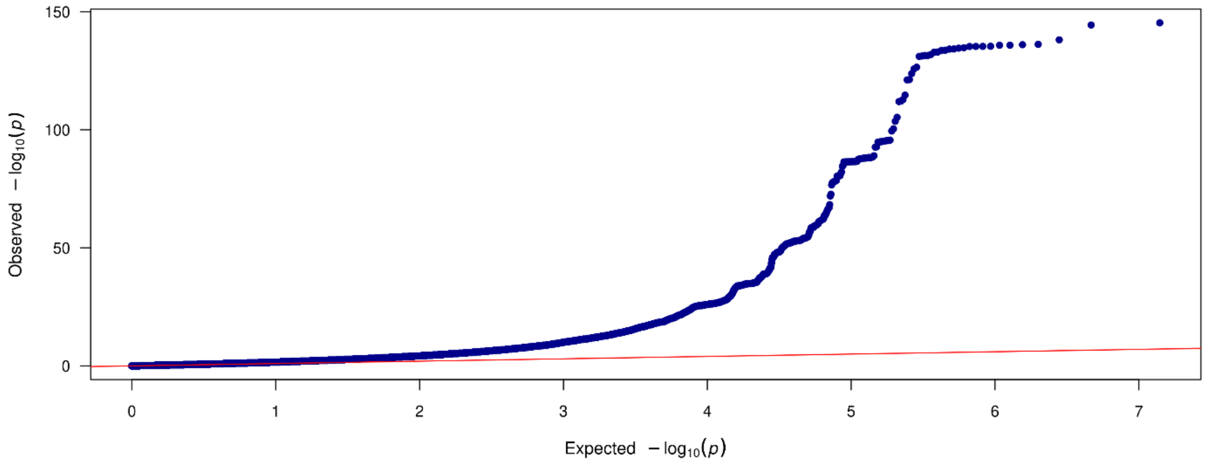

**Supplementary Figure 3. Manhattan and Quantile-Quantile (QQ) plot for Multivariate Male Adiposity GWAS.**

**a**, Manhattan plot showing GWAS results for the male adiposity factor plotted by chromosomal location. Chromosomal position is shown on the x-axis, with the y-axis displaying −log10(P) values from association tests between SNPs and the latent adiposity factor. **b,** Quantile–quantile (QQ) plot comparing observed versus expected −log10(P) under the null for the same GWAS.

**a**

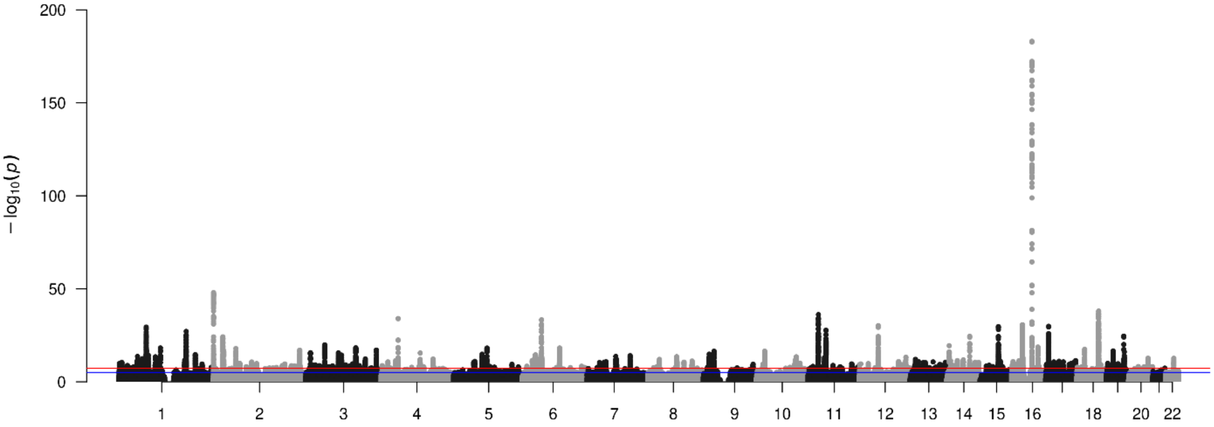

**b**

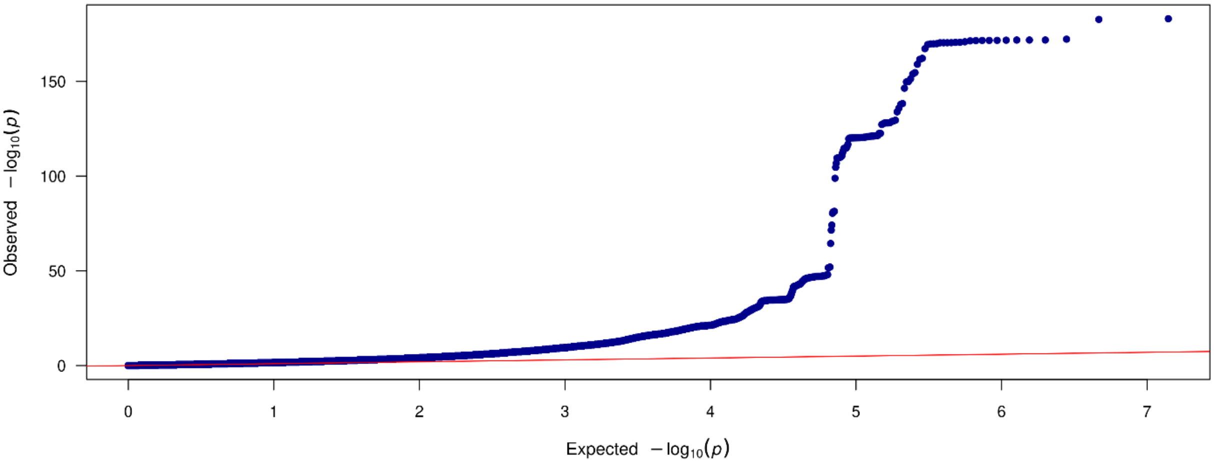

**Supplementary Figure 4. Manhattan and Quantile-Quantile (QQ) plot for heterogeneity test (i.e., QSNP) of Female Adiposity GWAS**

**a**, Manhattan plot of **Q_SNP** statistics from the Genomic SEM heterogeneity test for the female adiposity factor. The x-axis shows chromosomal position and the y-axis shows uncorrected −log10(P) from a $\chi^2$ test (df=2) of between-trait heterogeneity. **b,** QQ plot of the Q_SNP test, with expected versus observed uncorrected −log10(P) values for the female adiposity factor.

**a**

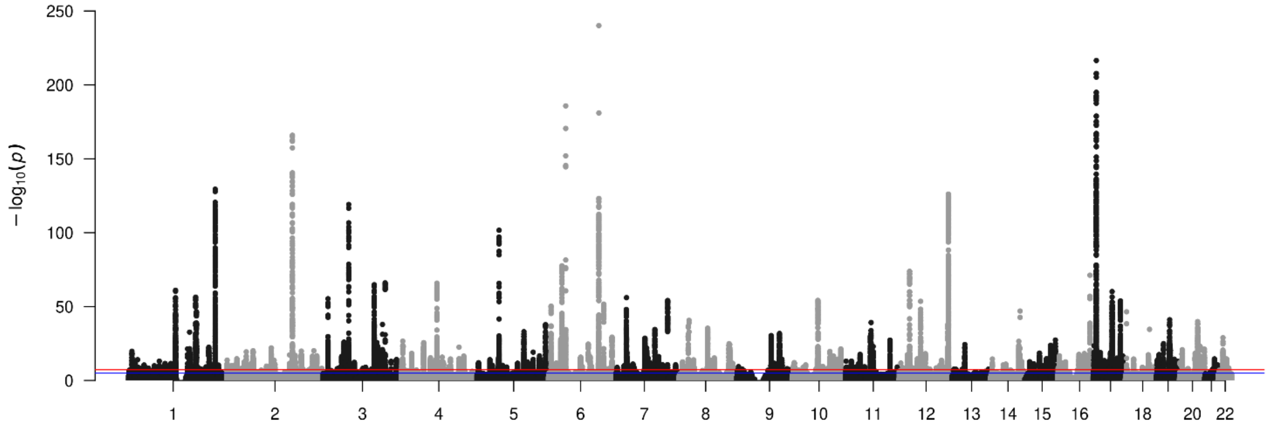

**b**

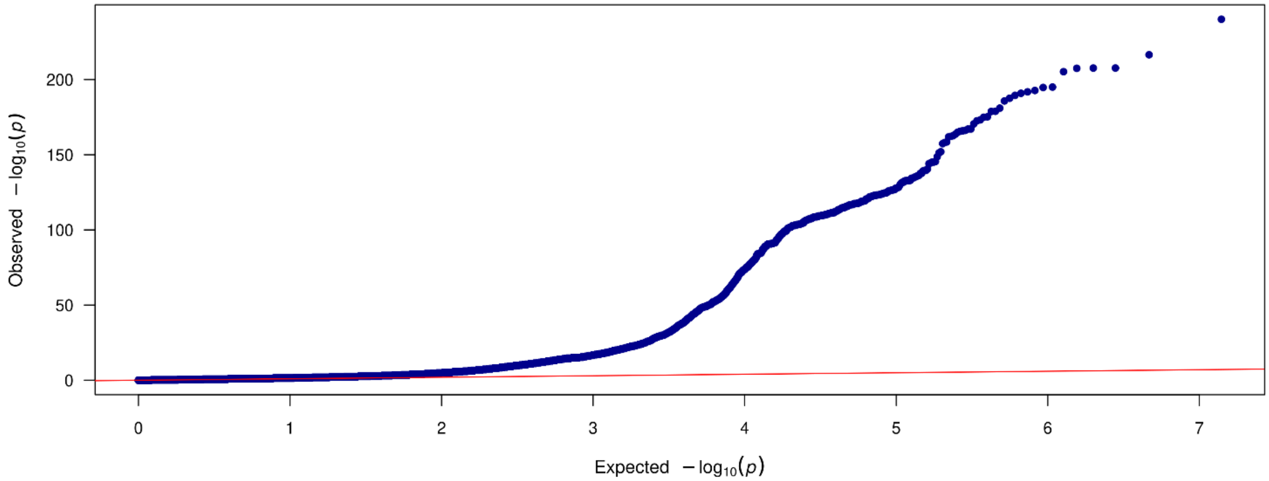

**Supplementary Figure 5. Manhattan and Quantile-Quantile (QQ) plot for heterogeneity test (i.e., QSNP) of Male Adiposity GWAS**

**a**, Manhattan plot of **Q_SNP** statistics from the Genomic SEM heterogeneity test for the male adiposity factor. The x-axis shows chromosomal position and the y-axis shows uncorrected −log10(P) from a $\chi^2$ test (df=2) of between-trait heterogeneity. **b,** QQ plot of the Q_SNP test, with expected versus observed uncorrected −log10(P) values for the male adiposity factor.

**a**

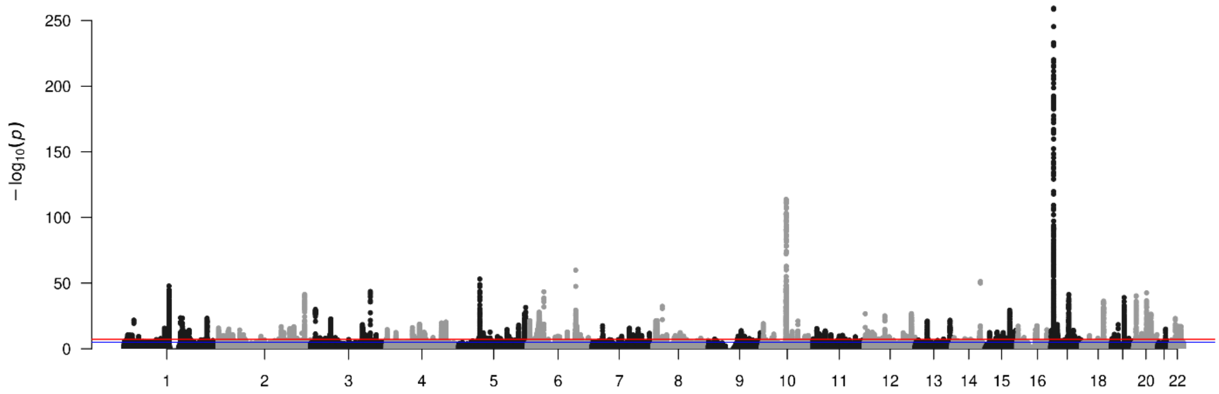

**b**

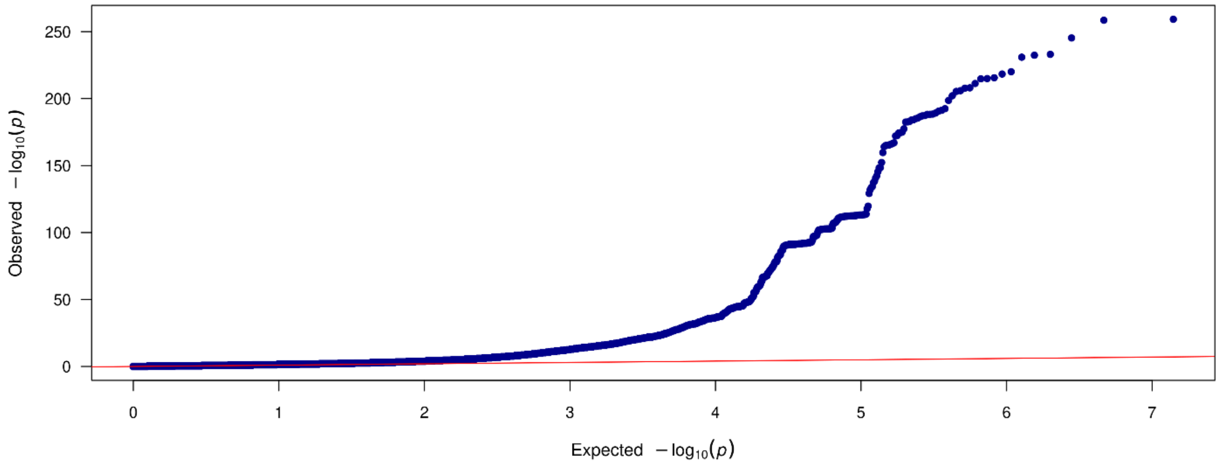

**Supplementary Figure 6. MiXeR-based partitioning of causal variants between sexes.**

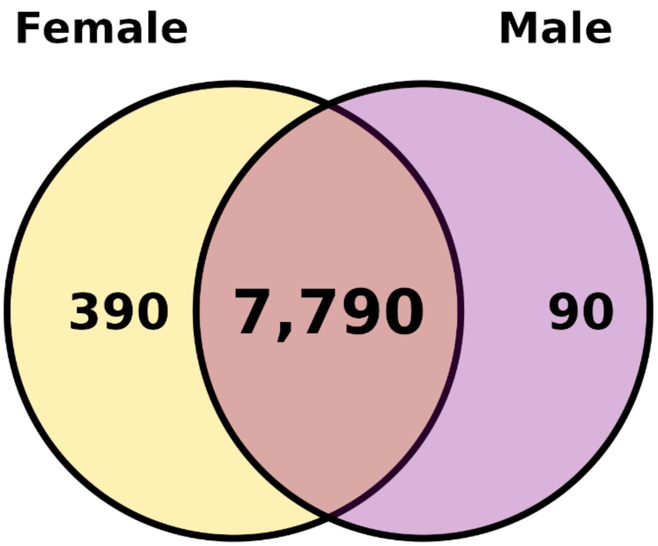

Venn diagram illustrating the overlap of inferred causal variants for the female and male adiposity factors. MiXeR estimated 8,180 causal variants in females and 7,880 in males, of which 7,790 were shared between sexes. The remaining variants comprised 390 female-specific and 90 male-specific components, representing the sex-dependent fraction of the polygenic architecture.

### Supplementary Figure 7. Miami plot of sex-stratified GWAS.

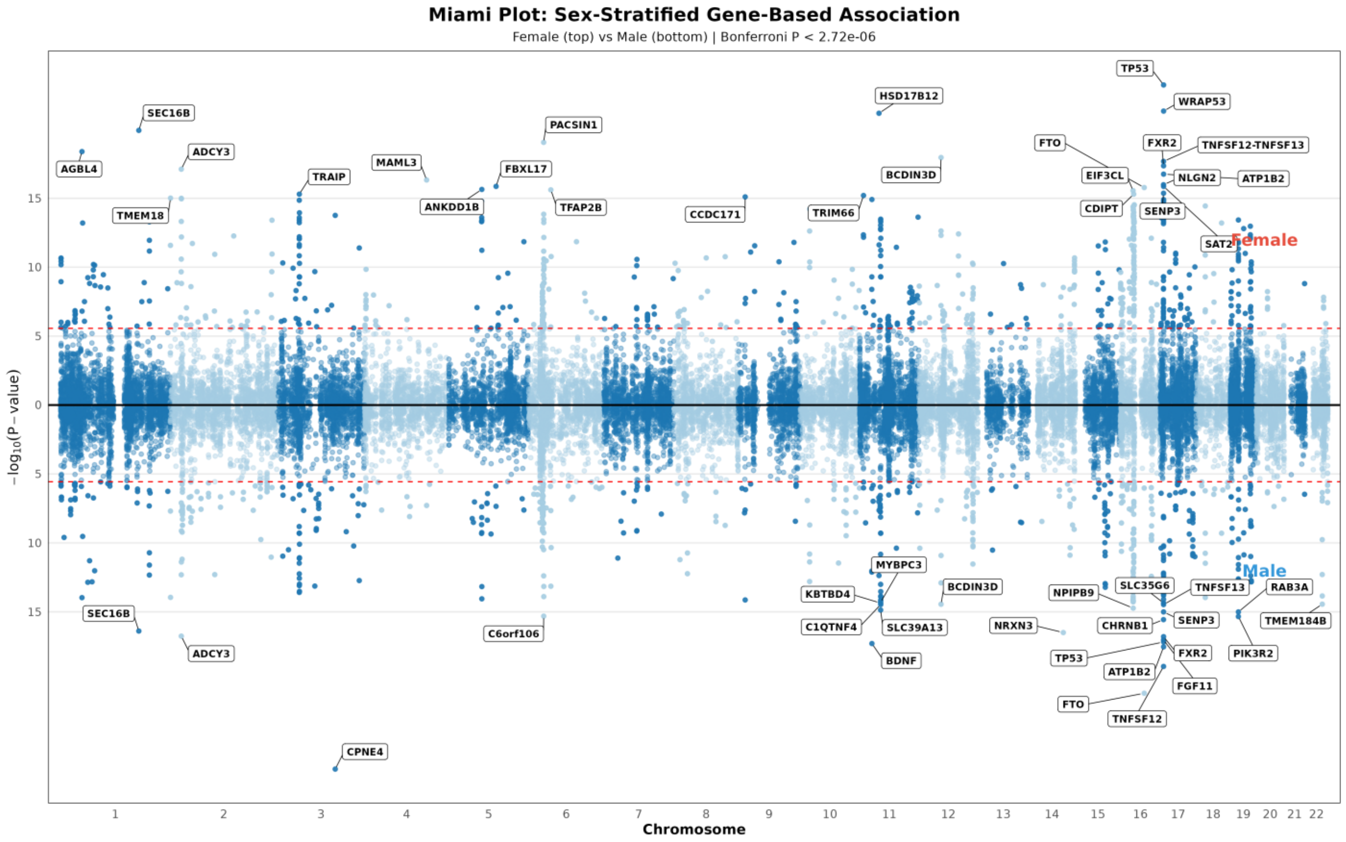

Miami plot comparing female (top) and male (bottom) MAGMA results across autosomes. The x-axis represents chromosomal position and the y-axis indicates −log10(P). The horizontal dashed line denotes the genome-wide significance threshold ($P < 5 \times 10^{-8}$).

**Supplementary Figure 8. Bayesian Colocalisation Posterior Probabilities for Endometrial Cancer and Adiposity-related Traits.**

*All locus-trait pairs shown demonstrated strong evidence of colocalisation (posterior probability ≥ 0.75) between endometrial cancer and at least one adiposity-related trait (BMI, WHR, BFP, ASAT, or VAT). The lead SNP, nearest gene, and adiposity-related trait are annotated on the left. Heatmap cells represent posterior probabilities for hypothesis 3 (distinct causal variants; PP.H3) and hypothesis 4 (shared causal variant; PP.H4) under 1MB and 200KB genomic windows.*

**Colocalisation Posterior Probabilities for Endometrial Cancer and Obesity-related Traits**

| SNP | Gene | Trait | PP.H3 1MB | PP.H4 1MB | PP.H3 200kb | PP.H4 200kb |
|---|---|---|---|---|---|---|
| rs17630235 | TRAFD1 | BMI | 0.0051 | 0.9948 | 0.0013 | 0.9987 |
| | | WHR | 0.0125 | 0.9574 | 0.0037 | 0.8922 |
| rs11065987 | SH2B3 | BMI | 0.4899 | 0.4878 | 0.1582 | 0.8338 |
| | | WHR | 0.0125 | 0.9680 | 0.0065 | 0.9677 |
| rs12602912 | BPTF | BMI | 0.1503 | 0.8489 | 0.1455 | 0.8538 |
| | | ASAT | 0.1525 | 0.6384 | 0.1283 | 0.6620 |
| rs4378452 | CUX2 | BMI | 0.9999 | 0.0001 | 0.0019 | 0.9627 |
| rs7127900 | TH | BMI | 0.0275 | 0.9718 | 0.0275 | 0.9718 |
| rs2891403 | RPH3A | BMI | 0.0153 | 0.9834 | 0.0159 | 0.9597 |
| rs2682911 | LINGO1 | BMI | 0.8949 | 0.0035 | 0.0839 | 0.8150 |
| rs4953042 | LRPPRC | BMI | 0.4932 | 0.2658 | 0.0865 | 0.6658 |
| rs8065496 | RAB11FIP4 | BMI | 0.2078 | 0.6656 | 0.1581 | 0.7251 |
| rs7498665 | SH2B1 | BMI | 0.1351 | 0.8154 | 0.1076 | 0.8414 |
| rs454388 | DLG2 | BMI | 0.3484 | 0.4017 | 0.0949 | 0.6933 |
| rs7357754 | GADD45G | BMI | 0.1440 | 0.5671 | 0.0816 | 0.6084 |
| rs2720681 | PTV1 | WHR | 0.2329 | 0.5054 | 0.0708 | 0.7820 |
| rs3796621 | IDUA | WHR | 0.1288 | 0.7810 | 0.1197 | 0.7891 |
| rs2235529 | WNT4 | WHR | 0.0863 | 0.6920 | 0.0503 | 0.7407 |
| rs9398794 | TRMT11 | WHR | 0.9999 | 0.0000 | 0.0297 | 0.9465 |
| rs370155 | H2AC14 | WHR | 0.2520 | 0.6981 | 0.0530 | 0.9149 |
| rs13214023 | ZKSCAN3 | WHR | 0.1882 | 0.7796 | 0.0315 | 0.9440 |
| rs17587597 | TRIM38 | WHR | 0.2021 | 0.7174 | 0.0950 | 0.8466 |
| rs13207082 | PRSS16 | WHR | 0.1237 | 0.7872 | 0.0112 | 0.8986 |
| rs4736359 | CYP11B1 | BFP | 0.2239 | 0.7616 | 0.1480 | 0.8371 |
| rs7969341 | SLCO1B1 | BFP | 0.0341 | 0.9458 | 0.0296 | 0.9501 |
| rs10953259 | BAIAP2L1 | BFP | 0.0957 | 0.7820 | 0.0256 | 0.8426 |
| rs10835917 | WT1 | VAT | 0.1283 | 0.8567 | 0.1254 | 0.8595 |

PP: 1.00, 0.75, 0.50, 0.25, 0.00

**Supplementary Figure 9. Multivariate GWAS-by-Subtraction GenomicSEM model**

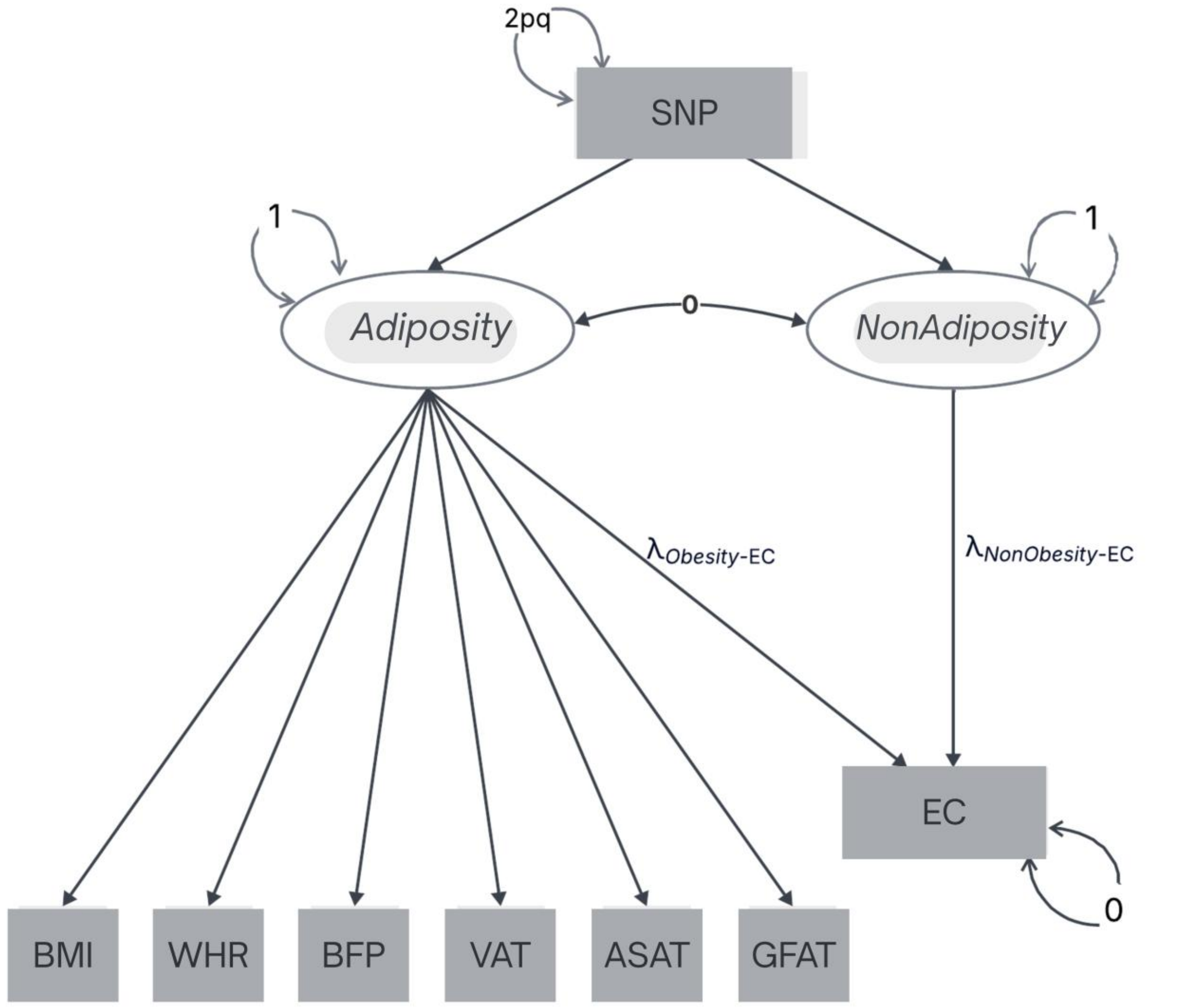

*GWAS-by-subtraction using GenomicSEM, partitioning SNP effects on endometrial cancer into adiposity-associated and adiposity-independent components via two orthogonal latent factors. Genetic effects across six adiposity traits (BMI, WHR, BFP, VAT, ASAT, GFAT) and endometrial cancer load onto a shared adiposity-factor, while the residual endometrial cancer loading defines the NonAdiposity pathway.*

**Supplementary Figure 10. Manhattan Plot of Non-Adiposity Component from GWAS-by-Subtraction Analysis**

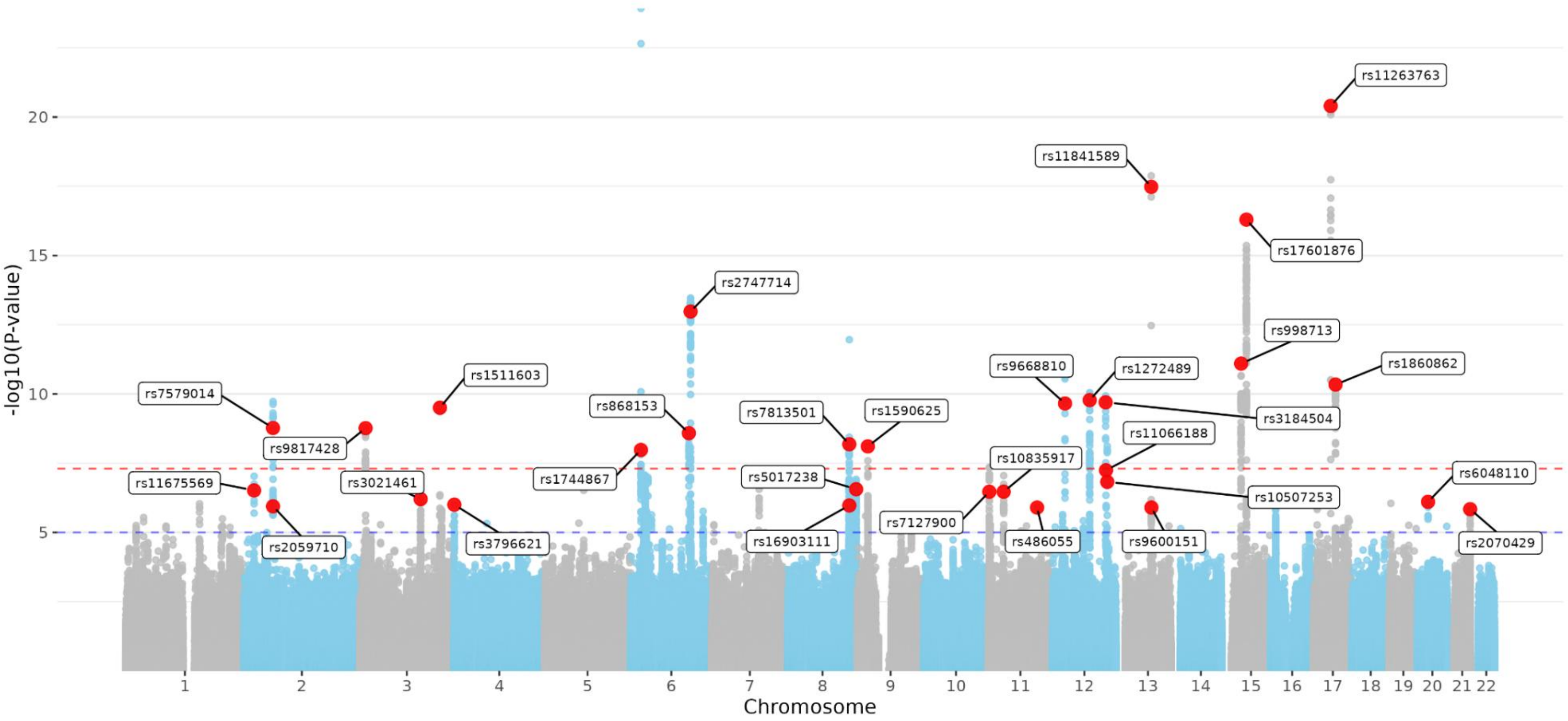

Manhattan plot showing genome-wide association results for the adiposity-independent component of endometrial cancer risk. X-axis shows chromosomal position; y-axis shows −log10(P). Red dashed line indicates genome-wide significance ($P < 5\times10^{-8}$). Top 30 lead SNPs are labeled.

**Supplementary Figure 11. Manhattan Plot of Adiposity Component from GWAS-by-Subtraction Analysis**

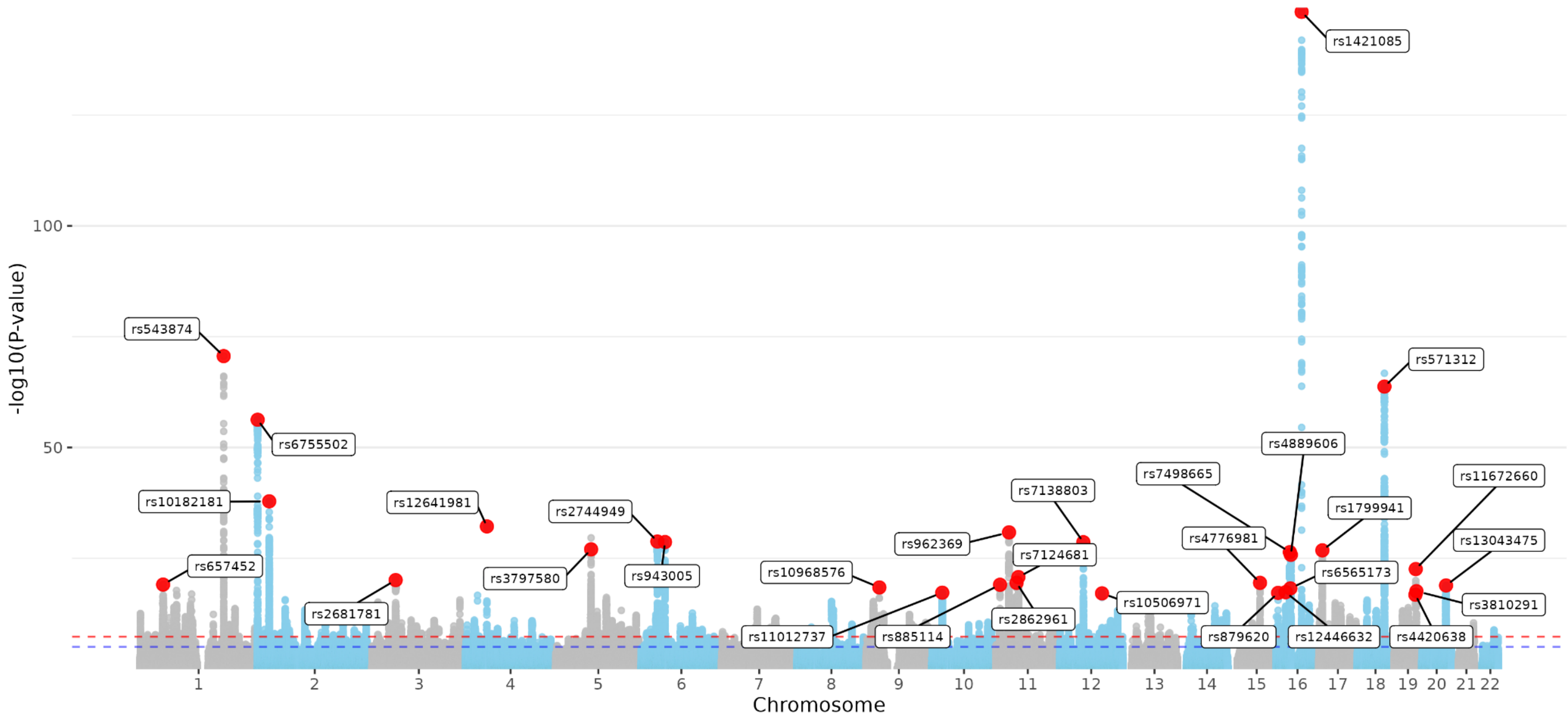

Manhattan plot showing genome-wide association results for the general adiposity component. X-axis shows chromosomal position; y-axis shows −log10(P). Red dashed line indicates genome-wide significance ($P < 5\times10^{-8}$). Top 30 lead SNPs are labeled.

**Supplementary Figure 12. Genetic correlations of NonAdiposity and Adiposity components with endometrial cancer-associated traits.**

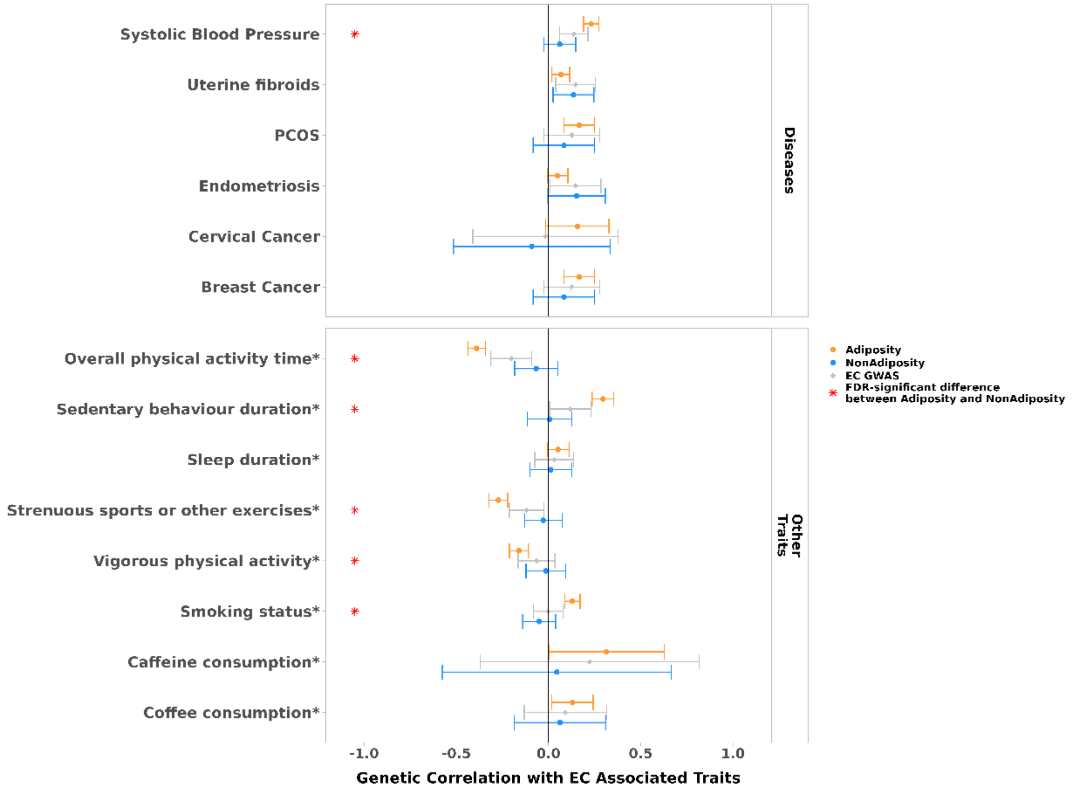

*Dots represent genetic correlations estimated using LDSC. Correlations with Adiposity are shown in orange, NonAdiposity components in blue, and endometrial cancer GWAS correlations in gray. Error bars indicate 95% confidence intervals. Red stars denote statistically significant differences (FDR-corrected P < 0.05, two-tailed test) between adiposity-dependent and adiposity-independent correlations, based on a Z-test that accounts for the standard errors of both estimates. FDR correction was applied across all correlations tested. Traits are grouped by disease-related and other endometrial cancer associated traits.*

**Supplementary Figure 13. Multivariate mediation model partitioning adiposity-mediated genetic effects on endometrial cancer risk.**

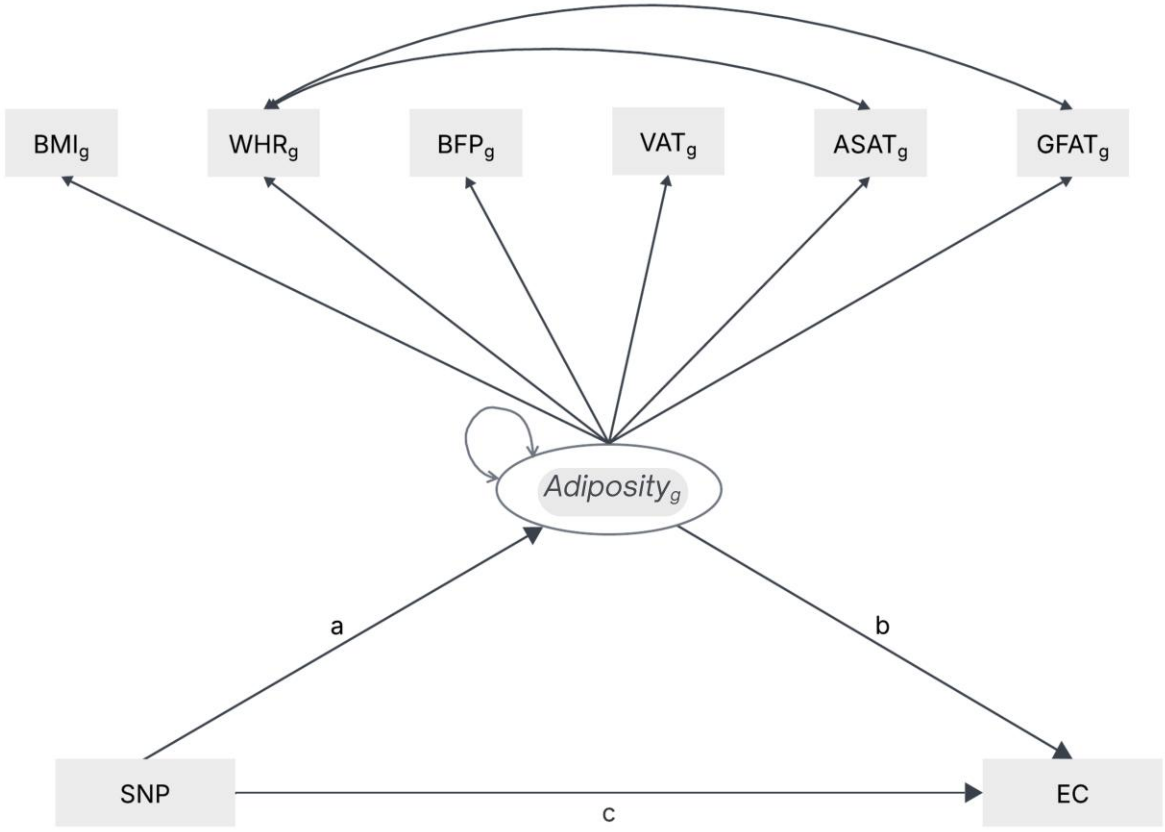

*Mediation model decomposing total SNP effects on endometrial cancer into direct (**c**) and indirect (**a** × **b**) components via adiposity. The indirect effect is calculated as the product of the SNP effect on adiposity (**a**) and the effect of adiposity on endometrial cancer (**b**). The total effect is: Total Effect = (c) + (a×b), and Proportion Mediated = Indirect Effect (a×b) / Total Effect (c+(a x b)).*

**Supplementary Figure 14. Pathway-specific signal characteristics and credible causal variant distribution across genomic regions**

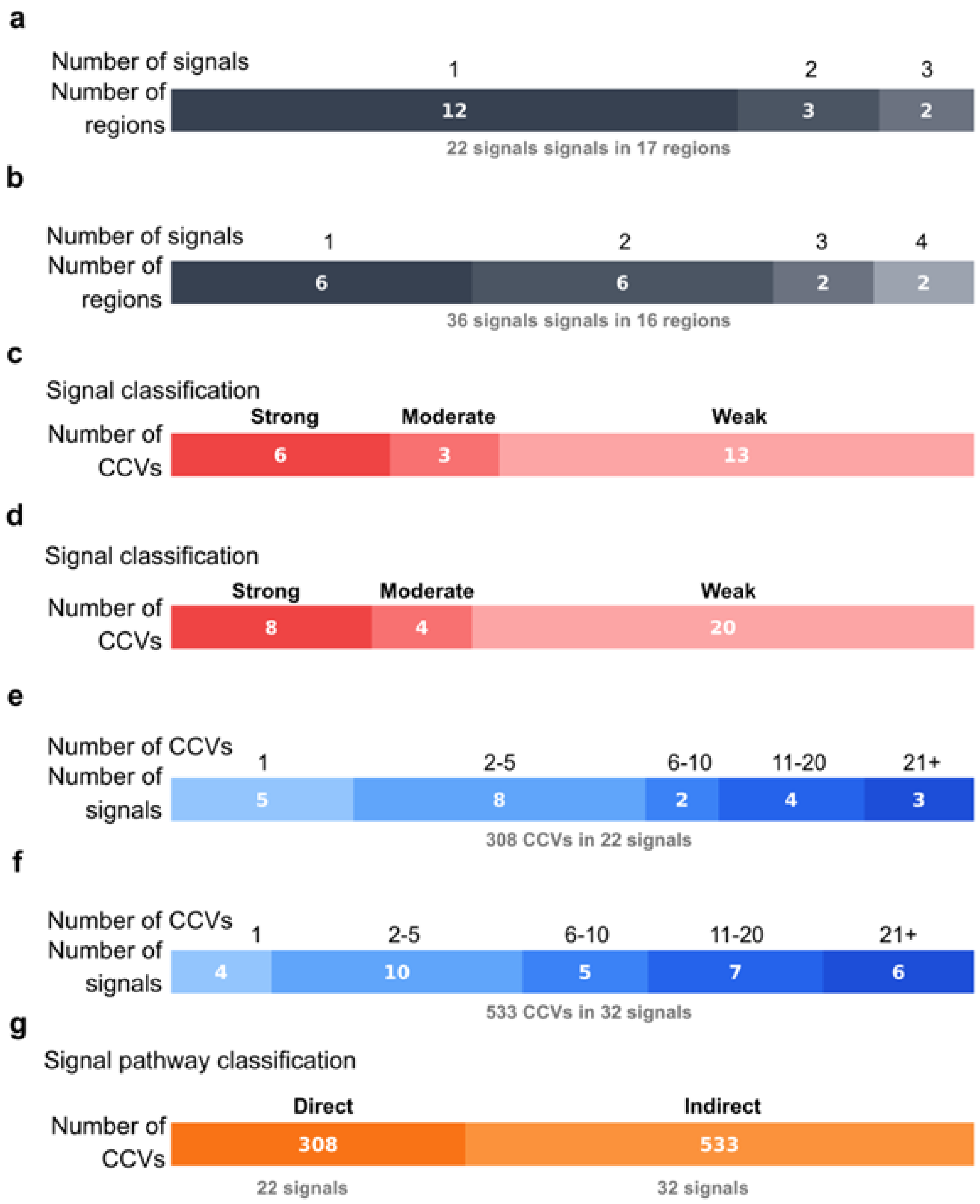

*Pathway analysis showing distribution of signals by number of genomic regions containing each signal count for direct (**a**) and indirect (**b**) pathways, signal classification by confidence levels (strong, moderate, weak) for direct (**c**) and indirect (**d**) pathways, and credible causal variant (CCV) count distribution per signal (**e**) Indirect pathway analysis with the same three-panel structure (**f**). Comparative summary of total CCVs between direct and indirect pathways (g).*
